# Strain-based biomarkers at the skin surface differentiate asymmetries in soft tissue mobility associated with myofascial pain

**DOI:** 10.1101/2024.12.18.24319267

**Authors:** Anika R. Kao, Terry M. Loghmani, Gregory J. Gerling

**Author notes:** **Correspondence:** Gregory Gerling, Olsson Hall 101D, 151 Engineer’s Way, Charlottesville, VA 22904.

## Abstract

Soft tissue manipulation is used widely to assess myofascial tissue qualitatively but lacks objective measures. To quantify the mobility of myofascial tissue, this effort derives optical biomarkers from the skin surface, as observed in the hands-on workflow of clinicians. Digital image correlation using three high-resolution cameras captures the cervicothoracic region as a clinician deeply engages and stretches the skin and myofascial tissue. Nineteen participants were positioned prone and marked with semi-permanent tattoos, optimized for tracking tissue without compromising its natural mechanics. Tissue mobility was then clinically assessed both bilaterally (left and right sides of body) and directionally (superior and inferior directions of pull). Eleven strain-based biomarkers were derived per tissue pull. With participants’ data aggregated, the sides of the body were indistinct, though pull in the superior direction was distinct from that in the inferior direction. Given substantial variance in the biomarkers’ absolute values between participants, we then evaluated each person individually. Therein, distinct tissue behaviors were observed. In particular, bilateral differences were identified in nine participants, eight of whom reported discrepancies in pain between their left and right sides, while directional distinctions were observed in sixteen participants, as expected given similar anatomical tissue structures between individuals. In our sample of participants, optical skin surface tracking and derived strain-based biomarkers identified asymmetrical distinctions in bilateral mobility, which correspond with self-reported pain. Such objective assessment of myofascial tissue stiffness is important in monitoring and treating chronic musculoskeletal pain, which afflicts half of the U.S. adult population.

**Highlights:**

- Clinicians use manual therapy to assess tissue mobility in musculoskeletal pain
- A hands-on clinical technique was paired with noninvasive 3D skin surface tracking
- Eleven strain-based biomarkers were developed to characterize soft tissue mobility
- Bilateral differences in mobility aligned with self-reported pain in most cases
- This approach offers a novel way to monitor musculoskeletal pain objectively

## 1. Introduction

Chronic musculoskeletal pain (MSK) afflicts up to half of the U.S. adult population, making it the most common reason for seeking massage therapy [1], [2], [3], [4]. For instance, the origins of neck pain often associated with headache [5], particularly tension headaches and migraines [6], are linked to tight muscles of the upper trapezius and levator scapula. Indeed, myofascial trigger points (MTrPs) – or tender spots within muscle or fascia associated with referred pain and restricted movement [7], [8], [9] – are commonly found in trapezius muscles. Despite the prevalence of myofascial trigger points and their associations with pain, muscle and fascial mobility and elasticity remain difficult to characterize [10].

Physical therapists use soft tissue manipulation (STM) with approximately 85% of their patients to examine and treat acute and chronic MSK [11], [12]. STM mechanotherapy imparts mechanical force through the intact surface of the body and can be delivered by hand contact alone or assisted with an instrument such as a rigid sphere or flat blade [13], [14], [15]. When performing STM, the clinician simultaneously assesses tissue tenderness, induration (hardness), restrictions, turgidity of inflammation, and mobility in multiple directions, using common palpation maneuvers including normal compression, distraction (pinch), and lateral stretch. STM stretch is characterized by the movement of tissue between layers, e.g., gliding of superficial over deeper tissue [16], [17]. While the magnitude, angles, and timing of forces delivered to and perceived from tissues seek to achieve distinct clinical effects, STM practice mostly relies on subjective, nonspecific, and qualitative descriptors [18], [19]. Indeed, despite patient self-reported outcomes and studies suggesting STM can relieve pain, including headache and associated cervical muscle tension [20], [21], [22], the mechanisms by which such mechanotherapies affect nervous, musculoskeletal, or integumentary systems remain unclear. In effort to understand the mechanisms by which such treatments produce clinical effects, a first step is developing quantitative biomarkers with robust signal-to-noise ratios, which can also support future clinical trials investigating their efficacy.

Numerous methods from ultrasound elastography to force-sensing devices have sought to characterize the viscoelastic properties of biological tissues *in vivo*. For instance, ultrasound-based methods measure displacement, shear wave differences, and microvascular changes to perturbations in muscle and nerve tissue [23], [24]. While recent studies have demonstrated the promise of ultrasound-measured biomarkers in discriminating pain states [25], intrinsic myofascial properties such as muscle anisotropy and a lack of well-defined imaging acquisition protocols challenge their wide-spread clinical deployment [26], [27]. Other imaging approaches, such as optical coherence tomography, can estimate tissue layer thickness and relative density, though only in planes a few millimeters from the skin surface [28], [29], [30]. Distinct from these imaging approaches, myotonometer devices use accelerometer recordings to assess tension, elasticity, and stiffness of myofascial tissues [31], [32], [33], while load cells generate force-displacement curves from which bulk stiffness may be derived [34], [35], [36]. Recent modeling efforts have sought to improve standardization and interpretability of these measurements through integrated approaches combining experimental data with finite element models [37]. However, these methods can only assess tissue properties over small contact areas, and small adjustments in probe angle and depth can lead to issues with repeatability and reproducibility.

In contrast, while optical cameras cannot penetrate muscle, observations at the skin surface can cover large areas and provide indirect information about underlying myofascial tissue mobility without impeding clinicians’ current workflow or requiring instrumented contacting devices. Although often in fields of study outside of soft tissue manipulation, large tissue areas have been imaged using depth and general-purpose RGB cameras [38], [39], [40], [41], [42], [43] with surface movements characterized using distinct analysis approaches. For instance, disparity map approaches effectively capture 3D surfaces at distinct time points but do not track movements of individual pixels between time points, and therefore cannot characterize strain or stretch [43]. In contrast, digital image correlation (DIC) approaches track unique pixel patterns over time, allowing for the evaluation of surface strain; and further, can stitch together multiple 3D surfaces to track large areas (tens of centimeters) and curvatures at high resolution, while avoiding occlusions [44], [45], [46], [47], [48], [29], [28], [49]. While DIC is used in many engineering domains, its application to *in vivo* biological tissue is emerging.

This study develops and evaluates a novel method for characterizing soft tissue mobility using optical measurements of skin surface displacement and strain. The primary goal is to derive quantitative biomarkers that align with clinical observations made during soft tissue assessments. To ensure clinical applicability and minimize disruption to existing workflows, we focused herein on the clinician’s use of STM stretch in the cervicothoracic region, a common site for headaches and associated myofascial trigger points [6]. While clinically referred to as ‘stretch,’ this maneuver involves a manual pull applied by the clinician to the tissue. This technique effectively captures multidirectional mobility and is readily assessed optically from the skin surface.

Our primary hypothesis is that these biomarkers will be sensitive enough to detect variations in underlying tissue mobility within individuals, and that these variations will correlate with self-reported pain levels. If successful, this study will provide objective tools to enhance precision and consistency in clinical practice, ultimately advancing the field toward evidence-based, personalized therapeutic interventions.

## 2. Methods

This work employs 3D-DIC to characterize tissue mobility as a clinician engages the skin and myofascial tissue in the cervicothoracic region. Custom methods are developed for this purpose, including skin speckling and imaging from three synchronized cameras to define eleven biomarkers, based on the 1^st^ and 2^nd^ principal strains and the magnitude of the clinician’s applied pull. The derived biomarkers are evaluated with nineteen participants to assess bilateral (left and right sides of body) and directional (superior and inferior directions of pull) distinctions in soft tissue mobility. The analysis of these biomarkers is compared to participant self-reported ratings, as the current standard to capture ground truth with respect to one’s level of pain [50], [51], [52].

### 2.1. Concept of analysis

This study was intentionally designed to preserve the clinical authenticity of manual soft tissue manipulation stretch, capturing the maneuver as it is naturally performed by an experienced practitioner. Rather than imposing a controlled mechanical input as in traditional material characterization, we employed a powerful, non-invasive imaging approach to quantify the skin surface’s response to unconstrained, clinician-applied loading. This observational, “fly on the wall” strategy enables extraction of quantitative biomarkers without disrupting the tactile and perceptual dynamics that are central to clinical practice. As a necessary tradeoff of this approach, force and force-rate were not directly measured in this study due to a fundamental methodological constraint: current force-sensing technologies require a physical barrier between the clinician and the participant. Inserting an instrumented interface or rigid component would fundamentally alter the nature of the maneuver, disrupting the tactile cues available to the practitioner and the perceptual experience of the participant. Prior studies have shown that even thin films placed on the skin can significantly reduce the perceived pleasantness of physical contact [53], highlighting the sensitivity of human touch to such disruptions. These considerations informed our decision to focus on the observable surface response, maintaining the ecological validity of the assessment while enabling quantitative analysis—an essential first step given limited mechanistic understanding of soft tissue evaluation in clinical practice.

### 2.2. STM stretch assessment

To standardize the STM stretch maneuver, the clinician manually applied a pull to the tissue with static force at a 45-degree angle horizontal to the myofascial plane, using a gradual 3 s ramp to maximum intensity. This maneuver was performed bilaterally (left and right sides of the body) and directionally (superior towards the neck and inferior away from neck) from a central reference point (e.g., tender spot) [17], [23] (Fig. 1(a)). Key factors of the clinical assessment include the spatial distance and velocity of force propagation from the point of pull application, the point of force magnitude and deformation at which discomfort (if any) is reached, the magnitude of force required to reach maximum myofascial stretch, and any identified restrictions or barriers to fascial motion (Fig. 1(c)). When in a healthy mobile state, superficial skin and subcutaneous tissues glide over deeper tissues, whereas tissues restricted by MTrPs exhibit resistance to the pull, clinically observed as a reduced distance of propagation (Fig. 1(d)).

**Fig. 1.**
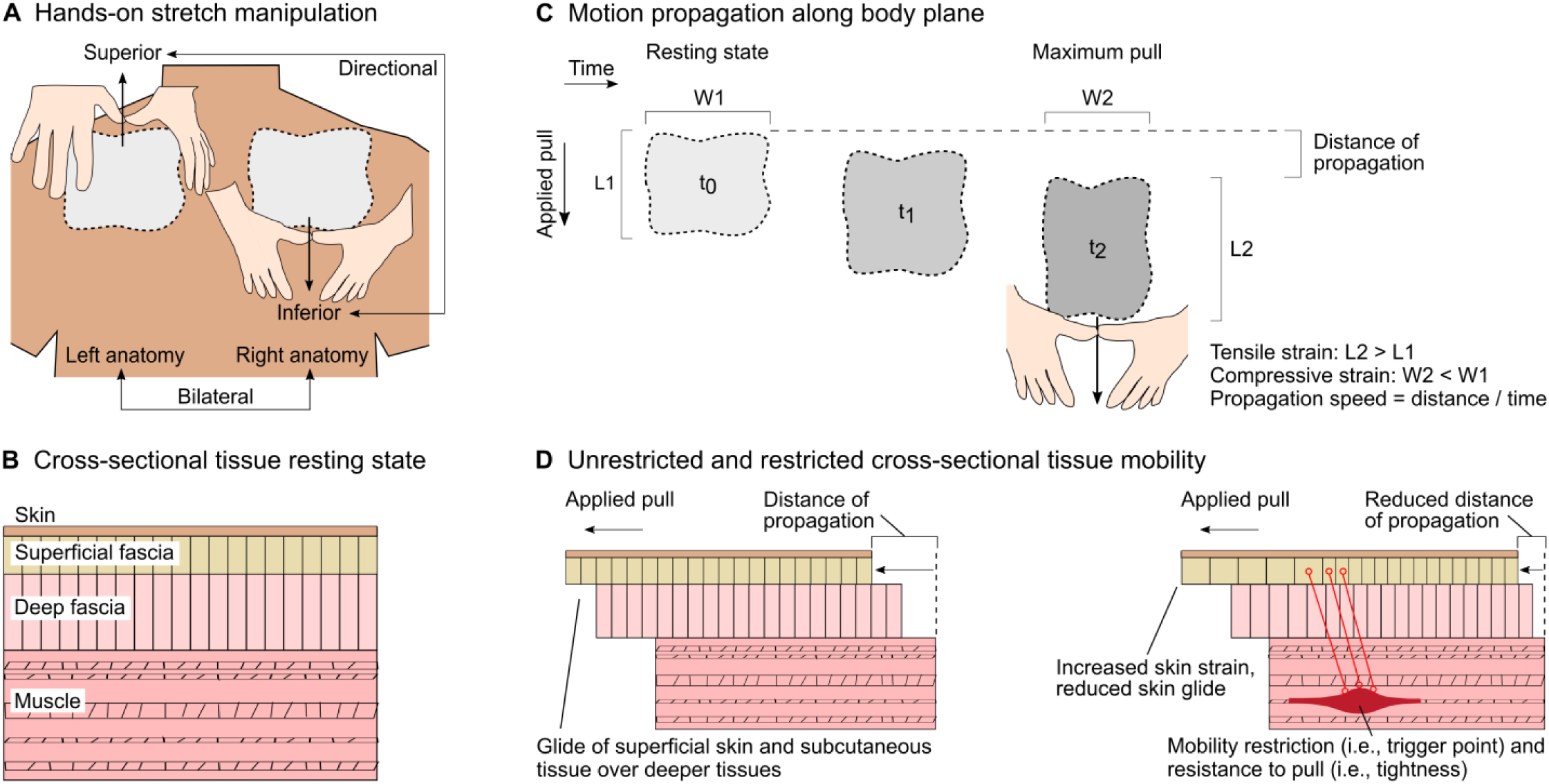
Translating clinical STM stretch assessment procedures and perceptual cues into quantitative analysis. (a) Schematic of STM stretch applied bilaterally (left and right sides of body) and directionally (superior and inferior directions of pull) in the cervicothoracic region. (b) Abstracted cross-sectional schematic of human tissue layers, including skin, superficial fascia, deep fascia, and muscle. (c) As the clinician increases force magnitude from resting state to maximum pull, the tissue not only expands vertically (tensile strain), but also contracts horizontally (compressive strain). (d) Comparative schematic of tissue respon se in healthy (left) and mobility restricted (right) conditions. While healthy tissue allows for superficial skin and subcutaneous tissue to glide over deeper tissues, tissue impaired with myofascial trigger points exhibits resistance to the pull, observed through reduced pull and propagation distances that a clinician can feel and often see.

To evaluate the consistency of clinician-applied input, a key assumption underlying our within-participant analysis of tissue response, we tracked the clinician’s fingers during three repeated trials on each side of a representative participant. Pull velocity was highly consistent within each side, with average rates of 14.8 ± 0.7 mm/s on the left and 20.9 ± 1.6 mm/s on the right (Supp. Fig. 1). Although velocities differed slightly between sides (6.1 mm/s), the influence of viscoelastic effects on strain output is expected to be negligible at such modest variations and timescales. In fact, prior work has shown that humans dynamically modulate displacement based on sensed mechanical impedance, adjusting their output using proprioceptive and cutaneous cues to maintain similar force levels when interacting with softer materials [54], [55]. This behavior highlights the clinician’s ability to finely tune manual input in response to subtle changes in tissue resistance, reflecting both tactile sensitivity and the learned capacity to adaptively take up slack based on years of hands-on experience.

### 2.3. Equipment setup and participant positioning

A portable massage table was set up whereby participants assumed a prone position to receive the STM assessment (Fig. 2(a)). The table’s face cradle was adjusted to ensure the neck was free of strain. A rotating arm used typically for computer monitors was positioned above the massage table (∼0.5 m) upon which were mounted, in a static and linear formation, three monocular cameras (12 MP, Raspberry Pi High Quality, UK) with wide angle lenses (6 mm Vilros, NJ, USA) connected to microcontrollers (Raspberry Pi Zero W boards, UK). The images from these cameras served as input into the image analysis pipeline and the rotating arm allowed participants to ascend the table while maintaining stereo camera alignment.

**Fig 2.**
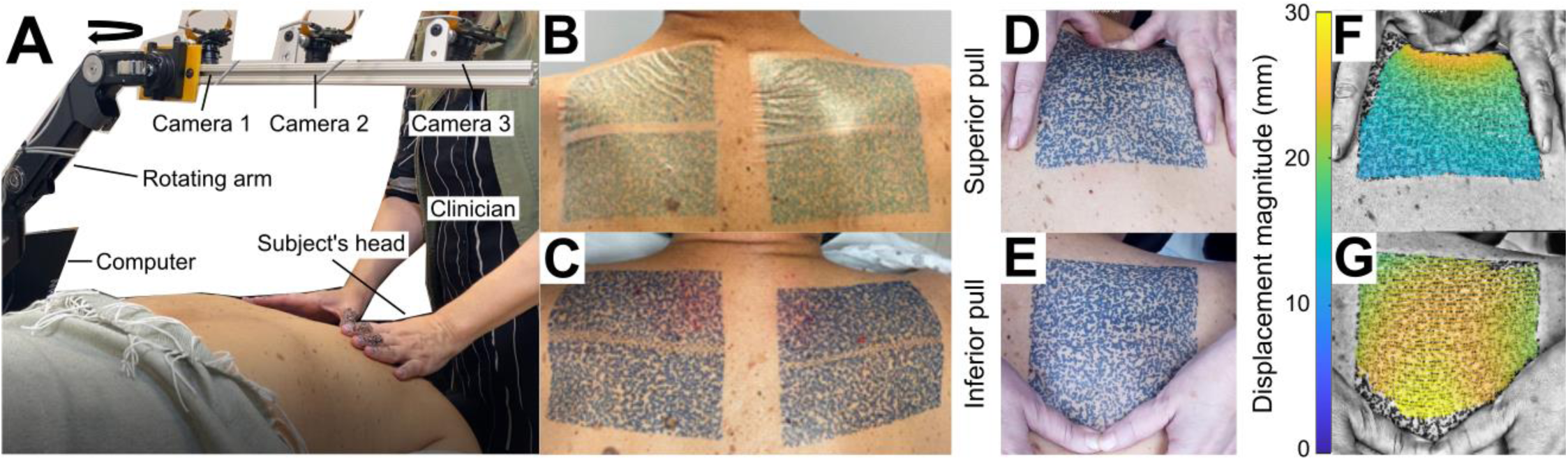
Equipment setup, speckling method, and STM stretch. (a) The clinician assesses the tissue mobility of the right cervicothoracic region of a participant. Participants assumed a standardized, prone position, with their arms at their sides (palms up) and feet resting on a bolster. The skin sur face is imaged with three overhead cameras mounted to a rotating arm. (b) Semi-permanent tattoo stickers are adhered, bilaterally, to the participant’s skin. Unspeckled bands between adjacent tattoo stickers are filled in with similar sized speckles using a semi-permanent tattoo pen. The tattoo stickers are peeled off after one hour. (c) After removal and a set time of at least 24 hours, the light blue speckles deepen to a dark blue color, enhancing contrast with the natural tone of the skin for improved tracking by 3D-DIC. The size, density, and randomness of the speckles inform the accuracy of the tracking and resolution of the resultant 3D point cloud. (d)-(e) Raw images showing pull applied manually by clinician’s fingers in the superior (towards neck) and inferior (away from neck) directions. (f)-(g) Colormap overlaid on raw image showing the change in skin surface displacement over the time duration of t = 0 → t = 2.7 s. Blue to yellow color hues indicate greater movement, including rigid body motion, in all three directions (X, Y, Z) as compared to the tissue’s resting state.

### 2.4. Participants

The experiment enrolled a total of 19 healthy participants (nine male, ten female, 34.1 ± 15.2 years of age, mean ± SD). All participants provided written consent, as approved by the University of Virginia Institutional Review Board of Social and Behavioral Sciences (Protocol #6201; Approved October 25^th^, 2023). This study was conducted as part of the clinical trial, “Optical Measurements of the Skin Surface to Infer Distinctions in Myofascial Tissue Stiffness (OptMeasSkin),” registered at ClinicalTrials.gov (ID: NCT06390085). All surfaces were sanitized regularly.

### 2.5. Semi-permanent speckling method

In using digital image correlation, attaining displacement fields of high spatial resolution depends on the absolute size and size consistency of applied speckles, the density and randomness of their pattern, and a high foreground-to-background contrast ratio with equal amounts of light and dark on the specimen surface. To meet these requirements and minimize the impact on natural skin mechanics, a semi-permanent ink tattoo method was developed. Four custom high-density speckle pattern stickers (∼1.5 mm speckle diam; 15.2 cm x 7.6 cm; InkBox, Toronto, Canada) were applied to the cervicothoracic region of each participant to create two regions of interest (15.2 cm x 15.2 cm; 231.0 cm^2^), one on the left side and one on the right side (Fig. 2(b)). The tattoo stickers remained in place for one hour, during which the ink reacted with the skin’s proteins to create a blue stain. Afterwards, the tattoos were removed, and over the course of 24 hours, the light blue speckles deepened to a dark blue, which enhanced contrast for optimal 3D-DIC tracking (Fig. 2(c)). The formula works on all skin tones. In particular, the darker the skin pigmentation, the darker the tattoo appears. The patterns remain visible for up to ten days, after which the skin regenerates and the color naturally fades.

### 2.6. Experimental procedure

To begin each participant’s session, the clinician conducted a qualitative intake assessment lasting ∼20 min. Participants provided demographic and medical history and reported initial pain levels on a 0-10 numeric rating scale (with 0 for no pain, 10 for one’s worst pain) for each of their left and right upper back regions. The self-reported pain ratings were used as a proxy assumption for perceived tissue stiffness, rather than clinician assessment, to eliminate any bias that might be introduced by our study procedural constraints (e.g., the clinician nearby hearing a participant give a pain rating out loud). Following the intake assessment, participants were positioned on a massage table in a standardized prone position, with their arms at their sides (palms up) and feet resting on a bolster. The cervicothoracic junction (C7/T1), superior medial border of the scapulae, and distal insertion of the levator scapulae were palpated and marked with a black dot, bilaterally. The STM stretch maneuver was applied in two directions (superior and inferior) first on the right side of the body, then on the left. Each maneuver was repeated three times for a total of three trials per direction per body side for a total of 12 stretch experiments per participant, over a session duration of ∼20 min.

Fig. 2 illustrates the clinical application of STM stretch in the superior (Fig. 2(d)) and inferior directions (Fig. 2(e)). Data for the example participant in Fig. 2 show the change in displacement at the time of maximum applied pull (Fig. 2(f) and (g)). For instance, in Fig. 2(f), the greatest displacement (29.5 mm) occurs in the direction of the applied pull, closest to the point of contact with the clinician’s finger. Additionally, the directional movement of the skin is distinctly different between stretch applied in superior and inferior pull directions.

### 2.7. Imaging approach using 3D digital image correlation

Digital image correlation is a non-contact, optical tracking technique that uses cross-correlation of pixel patterns from multiple stereo camera angles to produce displacement and strain fields [56], [57]. In this method, the image is divided into defined portions called subsets, which allow for localized measurements of movement and strain. Prior to data collection, a stereo camera calibration step determines each camera’s field of view and ensures adequate overlap. This calibration enables multiple 3D surfaces to be stitched together to avoid occlusions and accommodate highly curved surfaces. While a single camera captures planar displacement, a calibrated pair of cameras correlates 2D information to measure 3D deformation.

We used open-source software MultiDIC [56], built atop Ncorr [57], to capture 3D skin surface displacements, strain and stretch fields, etc. Video from each of the three cameras was captured synchronously by parallel computing at 30 frames per second in 1920 pixels by 1080 pixels resolution (∼5 pixels/mm) and compressed into H.264 video format. As each trial lasted approximately 30 s, after videos were converted to images (∼900 images), data were down sampled to 15 frames/s to reduce excessive processing time. Raw images in grayscale were input into the software for computation. Based on the diameter of ink speckles on the skin and the nature of the surface deformation resulting from the STM stretch, the subset radius and spacing were set to 20 pixels and 10 pixels, respectively, to optimize feature tracking and data resolution. The skin surface was tracked throughout the time-series of each applied pull and resultant measures included 3D displacement magnitude and direction and 1^st^ and 2^nd^ principal strain, which were the foundational data relied upon in developing the eleven biomarkers.

### 2.8. Biomarker development

Eleven biomarkers were developed to characterize the spatial and temporal aspects of soft tissue mobility, including initial resistance to the applied pull and peak deformation. The DIC methods enable precise tracking of each subset, allowing us to evaluate both the distance of movement (tissue glide) and the degree of deformation (tissue strain) within the speckled area. For unrestricted tissue, we anticipate cohesive displacement across the tracked area, with movement closely following the applied pull and minimal local deformation, resulting in higher values for biomarkers measuring tissue glide. Conversely, restricted tissue is expected to behave as if tethered, causing discontinuous movement and localized deformation, likely leading to elevated levels of tensile and compressive principal strain. The derivation of each biomarker is described in the following sections using an example inferior pull of the right anatomy for a representative participant (Fig. 3(a)).

**Fig. 3.**
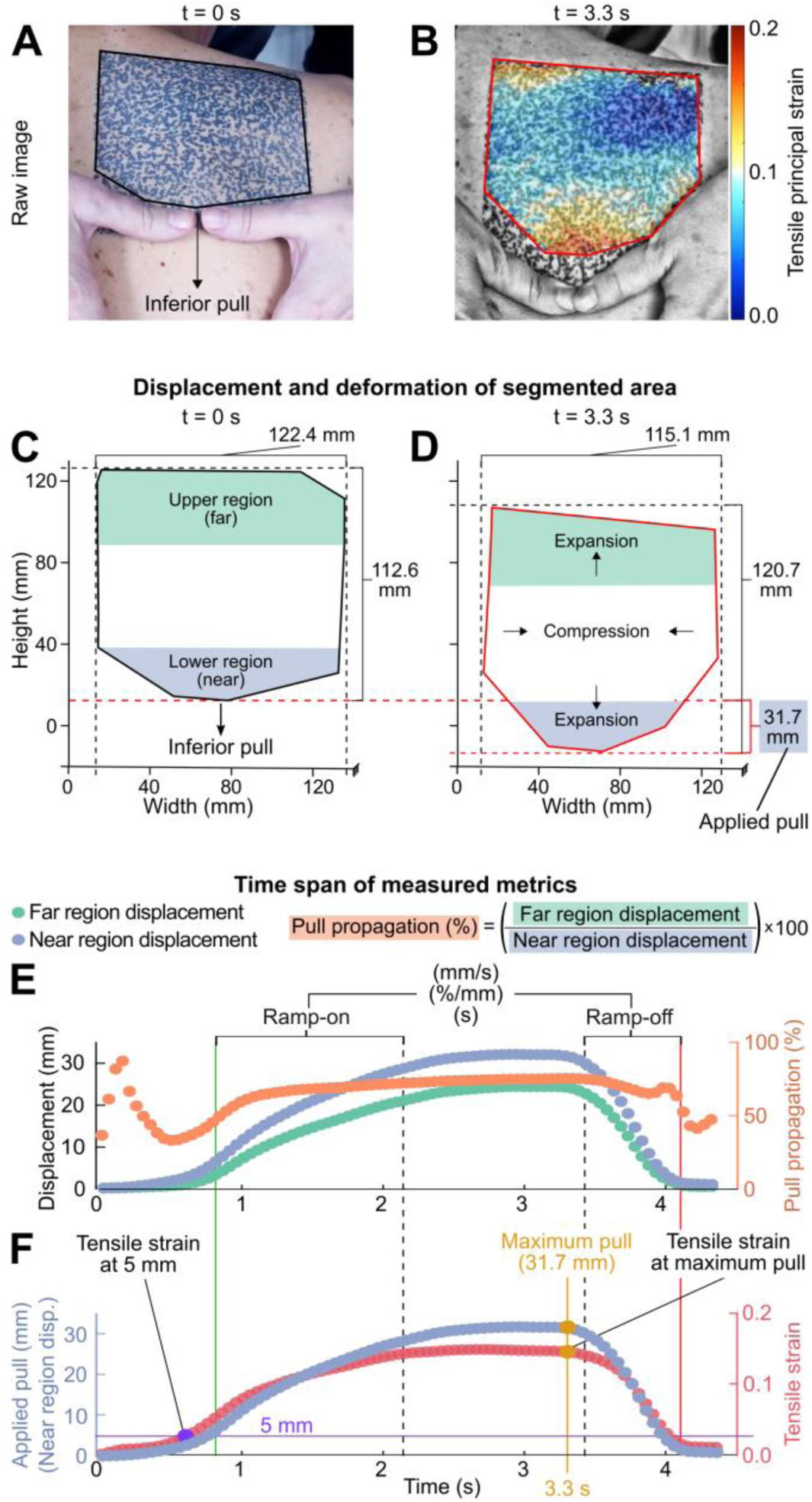
Derivation of biomarkers based on 3D skin surface measurements of STM stretch applied in the inferior direction. (a) Raw image of the tissue at initial state (t = 0 s) with the tracked area outlined in black. (b) Spatial distribution of tensile strain at maximum pull (t = 3.3 s), overlaid on the raw image. A tensile strain of 0.2 (red) indicates a 20% expansion relative to the initial state (blue). Notably, higher tensile strains, or greater expansion, are concentrated near the force application point. (c) Segmented area of the 3D point cloud tracked by DIC at initial state (t = 0 s), with upper and lower regions identified by their vertical position (upper 20% of points; lower 20% of points). (d) The segmented area at maximum pull (t = 3.3 s) demonstrating overall horizontal contraction (115.1 mm < 122.4 mm) and vertical expansion (120.7 mm > 112.6 mm). Applied pull, defined as the 95^th^ percentile of point-wise displacements in the region nearest to clinician contact (e.g., lower region for an inferior pull), measured 31.7 mm at t = 3.3 s. (e) Time series of displacements in the near (lower) and far (upper) regions, along with the pull propagation, defined as the percentage of far region displacement to near region displacement (e.g., upper region/lower region for an inferior pull). Both upper and lower regions exhibited increasing displacement with peak values at 3.3 s. However, due to the inferior pull direction, the lower region displayed greater maximum displacement (31.7 mm) compared to the upper region (24.2 mm). To standardize the analysis, each applied pull was divided into ramp-on, hold, and ramp-off phases. (f) Time series of the applied pull and tensile strain with strain at 5 mm and strain-to-pull at maximum pull labeled accordingly.

*Maximum Tensile and Compressive Strain:* The 1^st^ and 2^nd^ principal Lagrangian strains quantify the maximum tensile and maximum compressive deformation, respectively, of the skin surface in the tracked region. As illustrated in Fig. 3(b), a tensile strain of 0.2 (red hues) indicates a 20% expansion relative to the initial, undeformed state (blue hues), whereas a compressive strain of −0.2 represents a 20% contraction. Therefore, a lower value for compressive strain indicates greater contraction. Notably, higher tensile strains, or greater expansions, are concentrated near the application of pull, as indicated by the intense red color near the clinician’s fingers. At each time point, the 95^th^ percentile of the 1^st^ principal Lagrangian strain field is calculated following established methods [58], with the highest value throughout the pull reported as *maximum tensile strain*. Similarly, the 5^th^ percentile of the 2^nd^ principal Lagrangian strain field is measured at each time point, with the lowest value (most negative) recorded as *maximum compressive strain*, indicating maximum contraction.

Furthermore, the deformation of the entire tracked region can be evaluated by its change in width and length. Namely, at t = 0 s, the tracked region measured 122.4 mm x 112.6 mm (Fig. 3(c)), and at t = 3.3 s, the time of maximum pull, it measured 115.1 mm x 120.7 mm (Fig. 3(d)). Based on these measurements, there was a 6.0% decrease in overall width and a 7.2% increase in overall height. It is important to note, however, that these overall changes do not directly correspond to the strain values in Fig. 3(b), as the principal strains are calculated locally within each subset, rather than averaged over the entire area.

*Maximum Pull (mm) and Far Region Displacement (mm):* The tracked area (4,208 points) was divided into three regions based on vertical position: upper 20%, middle 60%, and lower 20% of points (Fig. 3(c)). The 95^th^ percentile of all point-wise displacements, including rigid body motion, in the upper and lower regions was recorded at each time point. As shown in Fig. 3(e), both regions exhibit an increase in displacement before peaking at 3.3 s. However, because this pull was applied in the inferior direction, the lower region shows greater overall displacement (31.7 mm) as compared to the upper region (24.2 mm). Therefore, the displacement over time in the region nearest to the location of pull delivery (e.g., the lower region for an inferior pull) is measured as the applied pull, with the *maximum pull (mm)* defined as the highest applied pull over time (Fig. 3(f)). Conversely, *far region displacement (mm)* is defined as the displacement in the region farthest from where the pull is delivered (e.g., upper region for an inferior pull) at a given time point.

*Ramp-On Pull Velocity (mm/s), Strain Gradient (1/mm), and Duration (s):* Each trial underwent a pre-processing pipeline to exclude frames captured before the start or after the end of the applied pull. Within this pipeline, trials were segmented into three sub-phases of ramp-on, hold, and ramp-off (Fig. 3(e)). Phase boundaries were determined using signal processing techniques, including derivative calculations and thresholding, to identify key inflection points in the pull and strain curves. The ramp-on phase is characterized by the *pull velocity (mm/s)*, *strain gradient (1/mm),* and *duration (s)* that capture the tissue’s initial resistance to deformation. Figs. 3(e)–(f) illustrate these three phases over an example pull. In these figures, we first encounter an increase in tensile strain during the ramp-on phase as the pull is applied, then a relatively stable hold phase as the pull application slows, and finally a steep decline during the ramp-off phase when the pull is released.

*Pull Propagation (%):* To assess the extent to which the applied pull propagates across the surface of the tissue, the percentage of far region displacement to near region displacement (e.g., upper region/lower region for an inferior pull) is calculated per time point (Fig. 3(e)). The *pull propagation (%)* biomarker is calculated as the average of this percentage across the hold phase and reflects a clinician’s visual assessment of the distance of propagation (Fig. 1(d)).

*Strain at 5 mm and Strain-to-Pull at Maximum Pull (1/mm):* To account for distinct tissue mobility between directions of pull and sides of the body, *tensile strain at 5 mm* of pull was defined per trial. As illustrated in Fig. 3(f), this biomarker facilitates directional and bilateral comparisons of initial tissue response at a standardized distance of pull. To characterize tissue behavior under peak load, the ratio of *tensile strain to applied pull (1/mm)* was calculated at the time point of maximum applied pull. By contrasting these biomarkers, we can differentiate the tissue’s response initially and under maximal load.

*Area Under the Curve (mm):* To evaluate tissue deformation over the entire trial while remaining time-independent, the integral of tensile strain (dimensionless) as a function of applied pull (mm) was calculated, yielding *area under the curve (mm)* (AUC). This biomarker captures the overall deformation experienced by the tissue across the applied pull, incorporating both the extent of tissue strain and the distance over which it occurs. Although AUC reflects the strain-displacement profile and may be sensitive to input velocity under certain conditions, pull velocity in this study was highly consistent, with only modest variation unlikely to induce meaningful viscoelastic effects.

### 2.9. Statistical analysis

To identify tissue mobility trends at the population level, we aggregated raw data from all participants, directions, and body sides. To analyze trends in pull direction and body side, we employed a linear mixed-effects (LME) model. This approach was chosen due to the inherent heterogeneity in tissue mobility across individuals and the repeated measurements within each pull direction and body side, which introduce non-independence in the data. The LME model accounts for these complexities by accommodating variability between individuals and dependences within the dataset, providing robust estimates of population-level trends. Before applying the model, we confirmed that its key assumptions—linearity, homoscedasticity, and normality of residuals—were satisfied through visual diagnostics, including residual plots and Q-Q plots. Statistical significance was assessed using an alpha threshold of 0.01.

Given the complex and multifactorial influence of body composition on tissue mechanics—including contributions from muscle mass, adiposity, age, and hydration—we focused on within-subject comparisons, assuming relatively consistent composition across sides and directions within the small test area (15.2 cm x 7.6 cm) [59], [60]. This design isolates tissue mobility differences within each participant and avoids comparisons across individuals. Although BMI was not controlled for, LME analysis with participant as a random effect revealed no significant correlation between BMI and any biomarker (p = 0.29-0.98, α = 0.01).

For individual level analysis, biomarker means were calculated across three trials for each pull direction and body side, resulting in four mean values per biomarker, i.e., left superior, left inferior, right superior, and right inferior. A Friedman test revealed a significant difference only in ramp-on velocity between trials 1 and 3 (p = 0.014), consistent with time-dependent viscoelastic properties of skin [61], while the remaining biomarkers showed no significant differences across trials. To compare means (e.g., left superior vs. left inferior), we evaluated three percent difference calculations.

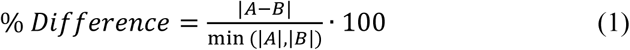

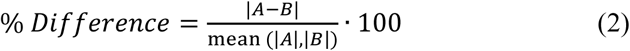

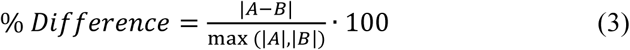

We conservatively chose to use (3), which resulted in four percent difference values per biomarker: left superior – left inferior, right superior – right inferior, left superior – right superior, and left inferior – right inferior. To classify whether a biomarker exhibited a significant differential response across pull direction or body side, we applied a 25% threshold, established in prior research [62], [63], [64]. Namely, any biomarker with a percent difference greater than or equal to 25% was considered significantly different. A total of 22 biomarkers were evaluated per comparison. For instance, a directional comparison included eleven biomarkers from the left side (superior versus inferior) and eleven from the right side (superior versus inferior). To assess whether the DIC method detected an overall difference, we applied another 25% threshold to the number of biomarkers showing significant differences. Participants were classified as having a directional or bilateral difference if six or more biomarkers (out of the 22) exceeded this threshold.

Self-reported pain ratings collected during the intake procedure served as a clinical assumption for associated aberrant tissue mobility. Participants with a difference of two or more points on the numeric pain rating scale (0-10) were considered to have underlying mobility asymmetries contributing to the reported bilateral pain differences. While pain ratings are subjective [50], [51], [52], studies have demonstrated correlations between self-reported pain and increased tissue stiffness in specific conditions [65], [66], suggesting that pain may offer clinically relevant insights into tissue behavior. Using these binary classifications, DIC-identified bilateral differences were compared with self-reported pain discrepancies to evaluate the clinical relevance of these novel tissue mobility measurements.

## 3. Results

### 3.1. Aggregate population trends

Across all participants, with pull directions and body sides aggregated, the clinician delivered a maximum pull of 30.3 ± 9.4 mm (median ± SD) (Fig. 4(a)), which produced 19.0 ± 9.1 mm of maximum far region displacement (Fig. 4(b)). The pull propagation biomarker, which measures the percentage of displacement that propagates across the tracked region, aligned with these values, measuring 65.9 ± 14.6% across the population (Supp. Fig. 2(a)). In response to the applied pull, maximum principal strains were 0.22 ± 0.07 in tension (Fig. 4(c)) and −0.14 ± 0.03 in compression (Fig. 4(d)). Since principal strain at the undeformed state (t = 0 s) is defined as zero, deviations from this baseline indicate the extent of tissue expansion or contraction. For instance, 0.22 signifies a 22% expansion along the primary loading direction, while −0.14 indicates a 14% transverse contraction. This pattern is consistent with expected tissue behavior under uniaxial tension, where lateral contraction arises as a result of longitudinal extension and is typically lower in magnitude due to approximate incompressibility.

**Fig. 4.**
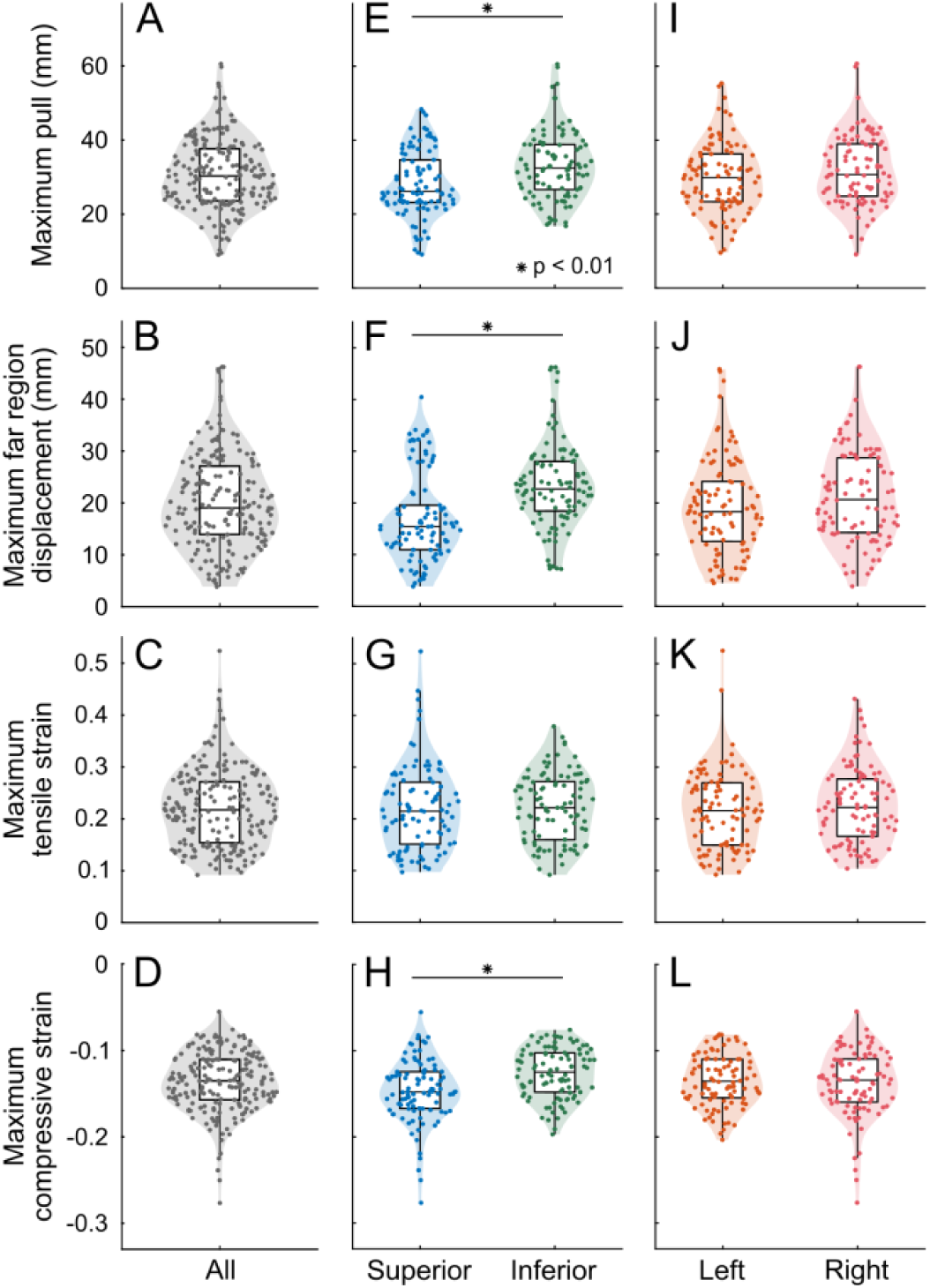
Aggregate population trends. (a)-(d) Biomarker distribution across all participants with sides and directions aggregated. In (a) the clinician delivered 30.3 ± 9.4 mm of maximum pull (median ± SD), which in (b) produced 19.0 ± 9.1 mm of maximum far region displacement. In response, in (c) tension, maximum principal strains measured 0.22 ± 0.07 and in (d) compression −0.14 ± 0.03. (e)-(h) Aggregate data separated by pull direction and analyzed using linear mixed-effects models (α = 0.01). Significant directional differences were observed in eight biomarkers (three shown: (e), (f), and (h)). Specifically, (e) maximum pull, (f) maximum far region displacement, and (h) maximum compressive strain were greater in the inferior direction. However, higher values of compressive strain in the inferior direction indicate more contraction in the superior direction. (i)-(l) Aggregate data separated by body side and analyzed using linear mixed-effects models (α = 0.01). No significant bilateral differences were found in aggregate for any of the eleven biomarkers.

Across all participants, with pull directions and body sides separated and analyzed using linear mixed-effects models (α = 0.01), significant pull direction differences were identified in eight of the eleven biomarkers (Fig. 4(e), (f), and (h); Supp. Fig. 2(h)-(j), (l), and (m)), though no significant body side differences were observed (Fig. 4(i)-(l); Supp. Fig. 2(o)-(u)). Specifically, median values of maximum pull (superior: 26.1 mm; inferior: 32.5 mm) (*t*(194) = 3.71, *p* < 0.01) (Fig. 4(e)), pull propagation (superior: 57.5%; inferior: 69.5%) (*t*(193) = 6.46, *p* < .01) (Supp. Fig. 2(h)), and ramp-on pull velocity (superior: 30.8 mm/s; inferior: 37.3 mm/s) (*t*(193) = 3.28*, p* < .01) (Supp. Fig. 2(i)) were all higher in the inferior direction. In contrast, biomarkers normalized by the magnitude of pull, including strain at 5 mm (superior: 0.09; inferior: 0.07) (*t*(194) = −6.52, *p* < .01) (Supp. Fig. 2(l)) and strain-to-pull at maximum pull (superior: 0.09 1/mm; inferior: 0.07 1/mm) (*t*(194) = −3.47, *p* < .01) (Supp. Fig. 2(m)), were consistently higher in the superior direction. In summary, these results suggest that tissue glide is greater given a pull in the inferior direction, while tissue strain is more pronounced given a pull in the superior direction under equivalent loading conditions.

### 3.2. Pull direction comparison for two representative participants

Given variability between participants, we evaluated each person individually and found that some exhibit clear mobility differences based on pull direction, while others do not. Fig. 5 highlights this contrast with a comparison of pull direction on the left anatomy for two representative participants, P12 and P18. For P12, the inferior (yellow) pull curve extends significantly further in both applied pull and tensile strain magnitude compared to the superior (blue) direction (Fig. 5(a)). In contrast, P18’s curves overlap almost entirely across the span of applied pull (Fig. 5(b)). Biomarker values were averaged across three trials, and mean values per pull direction and body side were compared using percent differences (section 2.8). Specifically, P12 exhibited a 52.4% difference in average maximum tensile strain (superior: 0.12; inferior: 0.26), compared to only a 4.7% difference for P18 (superior: 0.24; inferior: 0.25) (Fig. 5(c)). Similarly, P12 demonstrated a 61.6% difference in maximum pull (superior 11.0 mm; inferior: 28.6 mm), while P18 showed only a 1.9% difference (superior 40.3 mm; inferior: 39.5 mm) (Fig. 5(d)).

**Fig. 5.**
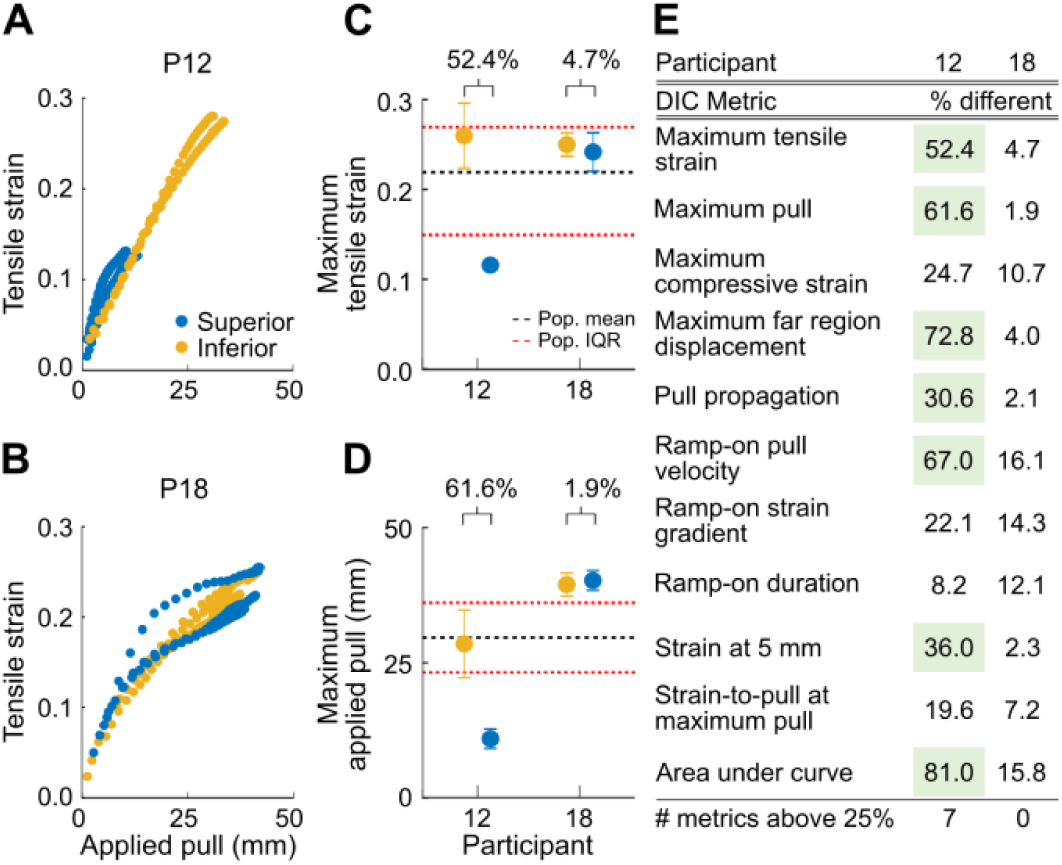
Pull direction comparison for two representative participants on the left anatomy. (a) Raw measurements of tensile strain and applied pull are shown for three trials of both superior (blue) and inferior (yellow) pull directions for participant 12 (P12). The inferior pull curve extends significantly further in both pull (28.6 mm) and strain magnitude (0.26) compared to the superior direction (11.0 mm; 0.12). (b) Applied pull to tensile strain curves for participant 18 (P18) appear nearly overlapping with similar values of maximum tensile strain (superior: 0.24; inferior: 0.25) and maximum pull (superior: 40.3 mm; inferior: 39.5 mm). (c) Percent difference between maximum tensile strain means in superior and inferior directions are higher in P12 (52.4%) compared to P18 (4.7%). Both of P18’s measurements were within the interquartile range (0.15 – 0.27) and close to the mean (0.22), whereas P12’s superior measurement fell outside of this range. (d) Maximum pull measurements followed a similar pattern, with P12 showing a 61.6% difference compared to only a 1.9% difference for P18. Both participants’ superior direction values fell outside of the population interquartile range (23.3 mm - 36.2 mm). (e) Similar analyses were conducted for the remaining nine biomarkers, with those flagged as directionally different by the criteria in section 2.8 highlighted in green. P18 showed zero biomarkers surpassing threshold, while P12 showed seven. These findings suggest that P12 exhibits directional mobility differences, unlike P18.

Analysis of the remaining nine biomarkers further supports this pattern: seven of P12’s biomarkers exceeded the directional difference threshold (section 2.8), while none of P18’s did (Fig. 5(e)). These findings suggest that P12 exhibits mobility differences based on pull direction, while P18 does not.

### 3.3. Bilateral comparison for two representative participants

Although no systematic bilateral trends were observed at the population level, individual-level analysis identified bilateral mobility differences in certain participants. Fig. 6 highlights this contrast with a bilateral comparison in the superior direction for two representative participants, P5 and P13. For P5, the applied pull to tensile strain curves nearly overlap (Fig. 6(a)), whereas P13 shows clear separation between the left (blue) and right (purple) sides (Fig. 6(b)). Quantitatively, P13 exhibited a 50.3% difference in maximum tensile strain (left: 0.24; right: 0.12) and a 31.5% difference in maximum pull (left: 15.1 mm; right: 22.1 mm). In comparison, P5 showed smaller differences of 13.3% and 17.7%, respectively (Fig. 6(c)-(d)).

**Fig. 6.**
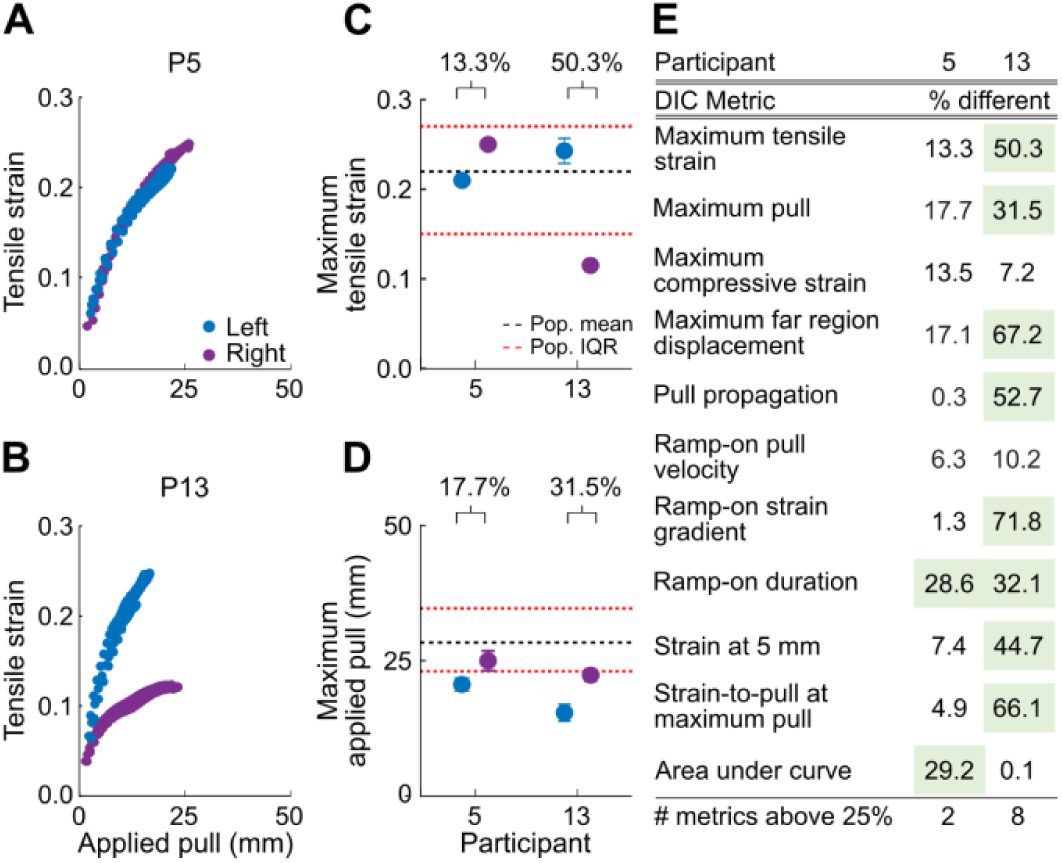
Bilateral comparison for two representative participants in the superior direction. (a) Raw measurements of tensile strain and applied pull are shown for three trials on both the left (blue) and right (purple) body sides of participant 5 (P5). Curves appear nearly overlapping with similar values of maximum tensile strain (left: 0.21; right: 0.25) and maximum pull (left: 20.4 mm; right: 24.8 mm). (b) Applied pull to tensile strain curves for participant 13 (P13) show clear separation between body sides with higher maximum tensile strain (0.24) but lower maximum pull (15.1 mm) on the left side compared to the right (0.12; 22.1 mm). (c) Percent difference between maximum tensile strain means on the left and right anatomy measured much higher in P13 (50.3%) compared to P5 (13.3%). Both of P5’s measurements were within the interquartile range (0.15 - 0.27) and close to the mean (0.22), whereas P13’s right side measurement fell outside of this range. (d) Maximum pull measurements followed a similar pattern, with P5 showing only a 17.7% difference compared to a 31.5% difference for P13. Both participants’ values on the left side fell outside of the population interquartile range (23.1 mm - 34.7 mm). (e) Similar analyses were conducted for the remaining nine biomarkers, with those flagged as bilaterally different by the criteria in section 2.8 highlighted in green. P5 showed only two biomarkers surpassing this threshold, while P13 showed eight. These findings suggest that P13 exhibits bilateral mobility differences, unlike P5.

Similar analyses across the remaining nine biomarkers (Fig. 6(e)) revealed that eight of P13’s biomarkers exceeded the threshold outlined in section 2.8, compared to only two for P5. These findings suggest that P13 exhibits bilateral mobility differences, while P5 does not.

### 3.4. Individual differences in pull direction across all participants

Sixteen participants (84.2%) exhibited differences in biomarkers between the pull directions. Comparisons of the effect of pull direction for two representative participants are illustrated in Fig. 5, highlighting patterns consistent with the broader dataset. The total number of biomarkers exceeding the threshold was counted per body side, and participants were classified as directionally different if they met the criteria outlined in section 2.8. These results, with detailed participant data, are provided in Table 1.

**Table 1.** Comparison of pull direction effects on the eleven biomarkers per participant. Percent differences ≥ 25% are highlighted in green and counted per body side. The combined total of biomarkers exceeding the threshold for both sides is tallied for each participant in the bottom row of the table. Participants who meet the criteria outlined in section 2.8 are highlighted in red and classified as directionally different based on the 3D-DIC analysis. In total, sixteen participants (84.2%) were identified as directionally different.

| Participant | 1 | 2 | 3 | 4 | 5 | 6 | 7 | 8 | 9 | 10 | 11 | 12 | 13 | 14 | 15 | 16 | 17 | 18 | 19 | Count of significant participants | Total count of significant participants |
| --- | --- | --- | --- | --- | --- | --- | --- | --- | --- | --- | --- | --- | --- | --- | --- | --- | --- | --- | --- | --- | --- |
| DIC Biomarker | Difference between superior and inferior measure on the **left** body side (%) |  |  |  |  |  |  |  |  |  |  |  |  |  |  |  |  |  |  |  |  |
| Maximum tensile strain | 21.3 | 39.5 | 44.0 | 75.3 | 10.0 | 5.4 | 25.1 | 28.6 | 5.4 | 3.7 | 34.7 | 52.4 | 12.4 | 15.8 | 25.1 | 33.2 | 28.5 | 4.7 | 29.9 | 11 | 19 |
| Maximum pull | 5.3 | 8.5 | 7.0 | 42.2 | 36.9 | 22.0 | 1.5 | 6.0 | 36.9 | 3.7 | 13.2 | 61.6 | 37.6 | 26.5 | 6.6 | 40.3 | 49.4 | 1.9 | 23.8 | 8 | 18 |
| Maximum compressive strain | 0.9 | 44.8 | 36.7 | 36.2 | 1.2 | 11.4 | 11.4 | 4.3 | 8.3 | 34.3 | 3.8 | 24.7 | 3.4 | 28.7 | 28.6 | 10.3 | 32.1 | 10.7 | 38.2 | 8 | 16 |
| Maximum far region displacement | 27.8 | 56.8 | 27.1 | 44.9 | 58.5 | 35.8 | 24.1 | 10.6 | 65.2 | 14.1 | 25.0 | 72.8 | 72.6 | 39.4 | 21.7 | 43.9 | 58.3 | 4.0 | 22.6 | 13 | 28 |
| Pull propagation | 34.4 | 54.2 | 31.8 | 68.8 | 35.2 | 19.0 | 22.2 | 18.1 | 45.4 | 11.1 | 35.8 | 30.6 | 57.3 | 18.1 | 16.5 | 5.8 | 18.6 | 2.1 | 1.8 | 9 | 15 |
| Ramp-on pull velocity | 23.9 | 27.2 | 24.5 | 17.6 | 39.1 | 14.9 | 2.0 | 5.9 | 55.2 | 23.5 | 8.7 | 67.0 | 24.2 | 19.5 | 34.3 | 24.6 | 55.5 | 16.1 | 7.5 | 6 | 19 |
| Ramp-on strain gradient | 6.5 | 47.3 | 45.5 | 71.9 | 29.5 | 18.0 | 41.7 | 31.8 | 39.5 | 1.5 | 29.4 | 22.1 | 47.0 | 4.0 | 21.8 | 9.4 | 42.8 | 14.3 | 24.2 | 10 | 15 |
| Ramp-on duration | 14.7 | 26.9 | 10.5 | 24.0 | 8.2 | 23.5 | 35.6 | 2.9 | 26.1 | 17.6 | 6.1 | 8.2 | 20.9 | 12.2 | 6.3 | 6.0 | 19.0 | 12.1 | 10.8 | 3 | 7 |
| Strain at 5 mm | 4.0 | 47.0 | 22.5 | 27.2 | 22.3 | 22.1 | 22.5 | 32.7 | 16.0 | 15.3 | 3.2 | 36.0 | 45.7 | 33.4 | 32.5 | 5.6 | 16.7 | 2.3 | 5.1 | 7 | 13 |
| Strain-to-pull at maximum pull | 26.1 | 44.9 | 40.1 | 57.3 | 30.0 | 17.3 | 34.2 | 24.8 | 34.2 | 1.8 | 25.6 | 19.6 | 45.6 | 12.6 | 30.1 | 8.7 | 33.2 | 7.2 | 15.9 | 11 | 16 |
| Area under curve | 34.6 | 19.2 | 53.0 | 61.6 | 41.1 | 34.1 | 8.5 | 27.0 | 41.4 | 5.4 | 31.5 | 81.0 | 36.7 | 31.2 | 10.2 | 56.8 | 63.1 | 15.8 | 52.9 | 14 | 27 |
| Count of significant biomarkers | 4 | 9 | 7 | 9 | 7 | 2 | 4 | 4 | 8 | 1 | 6 | 7 | 7 | 5 | 5 | 4 | 8 | 0 | 3 | Count of significant participants | Biomarkers ranked by participant count |
| DIC Biomarker | Difference between superior and inferior measure on the **right** body side (%) |  |  |  |  |  |  |  |  |  |  |  |  |  |  |  |  |  |  |  |  |
| Maximum tensile strain | 48.1 | 11.5 | 10.5 | 20.5 | 13.2 | 33.7 | 0.9 | 29.7 | 1.0 | 31.0 | 9.2 | 17.5 | 56.5 | 49.8 | 25.3 | 10.6 | 13.8 | 6.8 | 33.2 | 8 | Maximum far region displacement |
| Maximum pull | 29.8 | 15.6 | 5.4 | 2.7 | 33.1 | 34.6 | 0.2 | 37.2 | 33.9 | 19.2 | 7.7 | 52.0 | 47.7 | 41.7 | 18.6 | 15.3 | 43.6 | 3.9 | 32.2 | 10 | Area under curve |
| Maximum compressive strain | 16.1 | 6.9 | 6.1 | 8.0 | 5.6 | 30.0 | 6.0 | 30.3 | 0.9 | 18.8 | 41.2 | 14.0 | 38.7 | 32.0 | 7.2 | 31.7 | 42.1 | 22.3 | 50.4 | 8 | Maximum tensile strain |
| Maximum far region displacement | 38.7 | 32.6 | 18.1 | 32.8 | 53.3 | 40.8 | 10.3 | 53.3 | 59.8 | 17.1 | 44.5 | 71.3 | 42.2 | 48.8 | 39.6 | 32.9 | 51.9 | 8.9 | 27.0 | 15 | Ramp-on pull velocity |
| Pull propagation | 13.1 | 23.5 | 21.7 | 34.3 | 30.3 | 11.8 | 10.6 | 24.1 | 38.5 | 7.5 | 42.0 | 42.5 | 8.1 | 13.2 | 26.6 | 21.2 | 14.8 | 4.2 | 5.9 | 6 | Maximum pull |
| Ramp-on pull velocity | 52.7 | 36.0 | 18.9 | 34.3 | 38.4 | 41.8 | 10.2 | 4.3 | 44.7 | 18.4 | 32.9 | 60.5 | 46.0 | 35.6 | 25.0 | 3.4 | 39.1 | 8.8 | 32.8 | 13 | Maximum compressive strain |
| Ramp-on strain gradient | 8.7 | 11.6 | 13.0 | 17.0 | 26.5 | 10.2 | 0.3 | 24.0 | 37.8 | 16.1 | 24.7 | 45.1 | 23.9 | 23.2 | 41.3 | 16.6 | 38.3 | 0.5 | 1.1 | 5 | Strain-to-pull at maximum pull |
| Ramp-on duration | 9.4 | 28.6 | 36.7 | 24.0 | 15.9 | 7.1 | 7.4 | 47.6 | 4.4 | 12.5 | 17.5 | 5.6 | 3.8 | 1.3 | 15.5 | 15.8 | 29.6 | 12.7 | 7.8 | 4 | Pull propagation |
| Strain at 5 mm | 7.2 | 9.2 | 5.7 | 38.8 | 3.0 | 22.1 | 19.5 | 14.7 | 27.2 | 29.7 | 4.7 | 50.0 | 8.2 | 23.6 | 15.9 | 32.9 | 9.4 | 25.4 | 14.4 | 6 | Ramp-on strain gradient |
| Strain-to-pull at maximum pull | 26.3 | 3.9 | 7.2 | 18.3 | 23.1 | 1.1 | 0.3 | 19.6 | 34.6 | 14.4 | 17.0 | 43.2 | 17.1 | 13.5 | 39.5 | 23.8 | 37.9 | 2.4 | 0.7 | 5 | Strain at 5 mm |
| Area under curve | 64.1 | 48.0 | 0.0 | 25.0 | 34.3 | 54.2 | 9.3 | 55.3 | 35.4 | 47.6 | 4.0 | 55.9 | 73.4 | 54.8 | 7.5 | 1.1 | 62.4 | 22.8 | 51.0 | 13 | Ramp-on duration |
| Count of significant biomarkers | 6 | 4 | 1 | 5 | 6 | 6 | 0 | 6 | 8 | 3 | 4 | 8 | 6 | 6 | 6 | 3 | 8 | 1 | 6 | **Total** |  |
| Left and right body sides |  |  |  |  |  |  |  |  |  |  |  |  |  |  |  |  |  |  |  |  |  |
| Total count of significant biomarkers | 10 | 13 | 8 | 14 | 13 | 8 | 4 | 10 | 16 | 4 | 10 | 15 | 13 | 11 | 11 | 7 | 16 | 1 | 9 | 16 directionally different by DIC |  |

### 3.5. Individual differences in bilateral anatomy across all participants

Nine participants (47.4%) exhibited differences in biomarkers between body sides. Comparisons of the effect of body side for two representative participants are illustrated in Fig. 6. The total number of biomarkers exceeding the threshold was counted per pull direction, and participants were classified as bilaterally different if they met the criteria outlined in section 2.8. These results, with detailed participant data, are provided in Table 2.

**Table 2.** Comparison of body side effects on the eleven biomarkers per participant. Percent differences ≥ 25% are highlighted in green and counted per pull direction. The combined total of biomarkers exceeding the threshold for both directions is tallied for each participant in the bottom row of the table. Participants who meet the criteria outlined in section 2.8 are highlighted in red and classified as bilaterally different based on the 3D-DIC analysis. In total, nine participants (47.4%) were identified as bilaterally different.

| Participant | 1 | 2 | 3 | 4 | 5 | 6 | 7 | 8 | 9 | 10 | 11 | 12 | 13 | 14 | 15 | 16 | 17 | 18 | 19 | Count of significant participants | Total count of significant participants |
| --- | --- | --- | --- | --- | --- | --- | --- | --- | --- | --- | --- | --- | --- | --- | --- | --- | --- | --- | --- | --- | --- |
| DIC Biomarker | Difference between left and right measure in **superior** pull direction (%) |  |  |  |  |  |  |  |  |  |  |  |  |  |  |  |  |  |  | Count of significant participants | Total count of significant participants |
| Maximum tensile strain | 23.4 | 56.2 | 15.8 | 34.0 | 13.3 | 4.3 | 23.2 | 3.5 | 10.3 | 28.0 | 6.8 | 31.1 | 50.3 | 16.1 | 29.1 | 29.4 | 16.8 | 0.1 | 13.6 | 7 | 12 |
| Maximum pull | 6.0 | 28.7 | 3.9 | 7.9 | 17.7 | 14.8 | 1.2 | 34.5 | 12.8 | 24.8 | 13.1 | 8.8 | 31.5 | 14.7 | 4.9 | 2.3 | 14.2 | 4.5 | 5.1 | 3 | 8 |
| Maximum compressive strain | 7.6 | 20.0 | 10.7 | 49.0 | 13.5 | 5.9 | 8.1 | 26.8 | 6.6 | 1.2 | 37.8 | 20.1 | 7.2 | 12.1 | 13.0 | 1.5 | 2.9 | 24.1 | 13.3 | 3 | 7 |
| Maximum far region displacement | 16.1 | 4.7 | 30.2 | 46.4 | 17.1 | 0.2 | 15.6 | 61.4 | 19.2 | 40.7 | 26.0 | 2.4 | 67.2 | 19.6 | 7.8 | 16.6 | 13.8 | 11.3 | 7.2 | 6 | 12 |
| Pull propagation | 24.7 | 22.8 | 34.0 | 51.4 | 0.3 | 14.4 | 13.5 | 41.0 | 8.3 | 17.0 | 16.0 | 12.1 | 52.7 | 5.4 | 12.4 | 15.1 | 0.7 | 6.4 | 3.6 | 4 | 4 |
| Ramp-on pull velocity | 15.5 | 28.9 | 32.6 | 36.3 | 6.3 | 50.2 | 17.5 | 12.8 | 3.8 | 23.4 | 5.4 | 19.0 | 10.2 | 12.9 | 22.3 | 23.4 | 7.9 | 6.0 | 23.0 | 4 | 10 |
| Ramp-on strain gradient | 13.7 | 23.5 | 22.8 | 42.8 | 1.3 | 6.6 | 21.6 | 48.0 | 1.2 | 2.3 | 9.6 | 19.7 | 71.8 | 26.5 | 32.8 | 30.6 | 0.5 | 11.2 | 27.9 | 7 | 12 |
| Ramp-on duration | 9.4 | 5.0 | 36.7 | 0.0 | 28.6 | 44.3 | 30.9 | 44.4 | 6.5 | 3.9 | 13.6 | 3.6 | 32.1 | 6.5 | 11.3 | 1.6 | 5.6 | 26.1 | 14.3 | 7 | 10 |
| Strain at 5 mm | 1.1 | 49.8 | 2.9 | 10.5 | 7.4 | 1.3 | 8.2 | 21.8 | 1.9 | 14.3 | 1.3 | 26.9 | 44.7 | 7.6 | 5.8 | 23.1 | 1.3 | 11.8 | 21.1 | 3 | 4 |
| Strain-to-pull at maximum pull | 17.6 | 39.0 | 14.6 | 28.1 | 4.9 | 10.7 | 22.9 | 44.1 | 1.0 | 6.0 | 7.5 | 25.3 | 66.1 | 28.3 | 25.1 | 32.1 | 2.0 | 4.5 | 26.0 | 9 | 12 |
| Area under curve | 10.9 | 60.0 | 2.5 | 32.1 | 29.2 | 22.8 | 6.0 | 29.4 | 14.3 | 42.3 | 13.5 | 44.0 | 0.1 | 21.4 | 36.2 | 15.3 | 14.0 | 9.9 | 18.7 | 7 | 15 |
| Count of significant biomarkers | 0 | 6 | 4 | 8 | 2 | 2 | 1 | 8 | 0 | 3 | 2 | 4 | 8 | 2 | 4 | 3 | 0 | 1 | 2 | Count of significant participants | Biomarkers ranked by participant count |
| DIC Biomarker | Difference between left and right measure in **inferior** pull direction (%) |  |  |  |  |  |  |  |  |  |  |  |  |  |  |  |  |  |  | Count of significant participants | Biomarkers ranked by participant count |
| Maximum tensile strain | 13.9 | 18.2 | 40.4 | 53.1 | 16.4 | 26.7 | 3.5 | 5.0 | 4.2 | 7.6 | 22.8 | 16.2 | 23.3 | 28.9 | 29.0 | 15.5 | 0.2 | 11.1 | 17.6 | 5 | Area under curve |
| Maximum pull | 29.3 | 22.7 | 8.4 | 35.5 | 12.8 | 1.6 | 0.5 | 2.0 | 8.6 | 3.4 | 7.9 | 12.3 | 42.6 | 32.3 | 17.1 | 31.1 | 4.3 | 2.5 | 6.2 | 5 | Maximum tensile strain |
| Maximum compressive strain | 21.8 | 25.9 | 47.0 | 13.1 | 17.4 | 16.0 | 9.3 | 0.4 | 1.0 | 20.1 | 9.0 | 18.9 | 41.1 | 8.0 | 11.5 | 25.0 | 12.1 | 12.8 | 7.5 | 4 | Maximum far region displacement |
| Maximum far region displacement | 28.8 | 38.9 | 14.5 | 34.5 | 6.6 | 7.6 | 0.3 | 7.5 | 6.7 | 16.8 | 0.0 | 7.5 | 31.0 | 32.2 | 16.4 | 30.2 | 0.7 | 6.5 | 1.7 | 6 | Ramp-on strain gradient |
| Pull propagation | 0.2 | 22.4 | 19.2 | 2.3 | 7.3 | 6.8 | 0.6 | 5.1 | 3.0 | 13.6 | 7.1 | 5.7 | 17.1 | 0.3 | 0.4 | 1.4 | 3.9 | 4.4 | 7.6 | 0 | Strain-to-pull at maximum pull |
| Ramp-on pull velocity | 26.4 | 19.2 | 9.1 | 49.2 | 7.4 | 27.2 | 24.4 | 3.2 | 15.8 | 18.5 | 22.3 | 3.0 | 36.0 | 30.4 | 11.4 | 44.2 | 20.7 | 2.2 | 5.7 | 6 | Ramp-on pull velocity |
| Ramp-on strain gradient | 11.6 | 22.1 | 38.6 | 40.8 | 5.4 | 31.3 | 25.8 | 0.3 | 1.4 | 15.5 | 15.2 | 12.2 | 30.1 | 0.3 | 10.5 | 24.6 | 6.9 | 3.0 | 5.9 | 5 | Ramp-on duration |
| Ramp-on duration | 14.7 | 7.1 | 10.5 | 0.0 | 7.5 | 21.5 | 13.7 | 2.9 | 24.4 | 25.0 | 24.2 | 16.4 | 10.7 | 4.9 | 1.7 | 11.8 | 39.7 | 26.6 | 4.1 | 3 | Maximum pull |
| Strain at 5 mm | 11.9 | 14.0 | 29.1 | 6.1 | 18.6 | 1.3 | 4.6 | 0.7 | 15.0 | 3.3 | 2.9 | 6.6 | 9.9 | 19.4 | 14.8 | 7.7 | 9.2 | 17.4 | 12.5 | 1 | Maximum compressive stretch |
| Strain-to-pull at maximum pull | 17.4 | 6.0 | 34.9 | 27.3 | 4.3 | 25.4 | 14.9 | 7.4 | 1.6 | 10.5 | 17.0 | 5.4 | 24.8 | 5.1 | 13.5 | 18.7 | 5.0 | 13.5 | 12.5 | 3 | Pull propagation |
| Area under curve | 38.4 | 37.8 | 51.8 | 65.2 | 21.1 | 10.0 | 6.8 | 13.2 | 5.5 | 3.9 | 24.1 | 23.0 | 58.0 | 48.4 | 38.1 | 49.5 | 12.2 | 1.7 | 15.5 | 8 | Strain at 5 mm |
| Count of significant biomarkers | 4 | 3 | 6 | 7 | 0 | 4 | 1 | 0 | 0 | 1 | 0 | 0 | 6 | 5 | 2 | 5 | 1 | 1 | 0 | **Total** |  |
| Superior and inferior pull directions |  |  |  |  |  |  |  |  |  |  |  |  |  |  |  |  |  |  |  |  |  |
| Total count of significant biomarkers | 4 | 9 | 10 | 15 | 2 | 6 | 2 | 8 | 0 | 4 | 2 | 4 | 14 | 7 | 6 | 8 | 1 | 2 | 2 | 9 bilaterally different by DIC |  |

### 3.6. Comparison of DIC-identified and self-reported bilateral differences

Bilateral differences identified through DIC analysis aligned with self-reported bilateral pain discrepancies in 84.2% of participants (16 out of 19). Fig. 7 shows the number of DIC biomarkers exceeding the threshold for each participant, with the dashed line indicating the cutoff necessary for an overall bilateral difference (section 2.8). Filled markers represent participants who reported bilateral pain discrepancies, while open markers indicate those who did not. Overall, the DIC method identified nine participants as bilaterally different, eight of whom self-reported bilateral pain discrepancies. Conversely, of the ten participants not identified as bilaterally different by DIC, eight also did not report bilateral pain discrepancies.

**Fig. 7.**
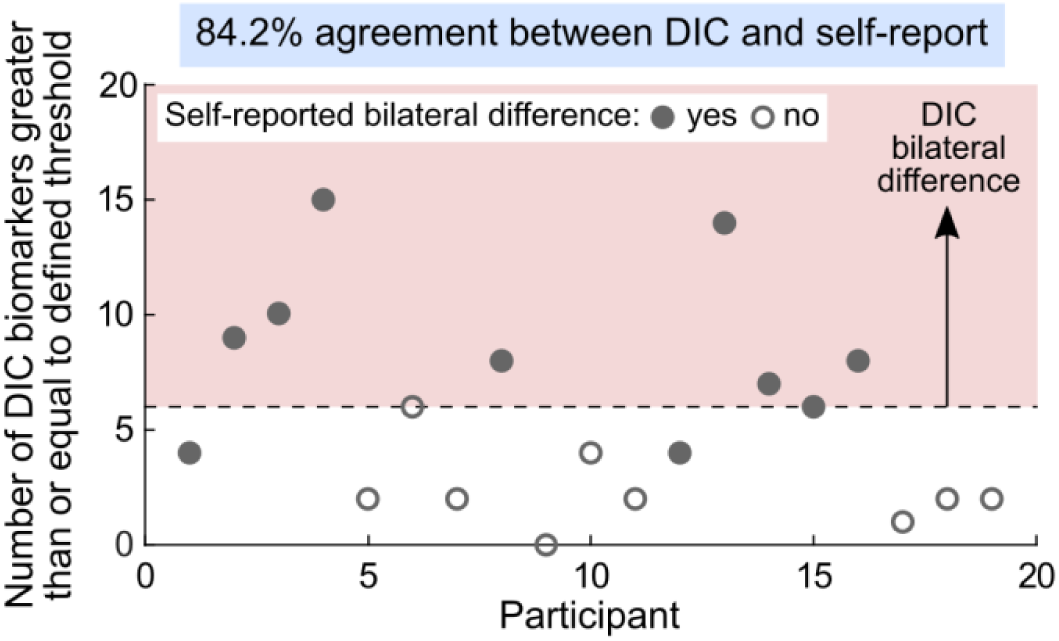
Comparison of DIC-identified and self-reported bilateral differences. The number of DIC biomarkers exceeding threshold is shown per participant, with the dashed line indicating the cutoff necessary for an overall bilateral difference (section 2.8). Filled markers represent participants who reported bilaterally different pain while open markers indicate those who did not. In total, the DIC method identified nine participants as bilaterally different, eight of whom self-reported bilateral pain discrepancies. Ten participants were not identified by DIC as bilaterally different, and eight of these did not report bilateral pain discrepancies. Overall, DIC-identified bilateral differences aligned with self-reported bilateral pain discrepancies in 84.2% of participants.

## 4. Discussion

Chronic musculoskeletal pain affects over 1.71 billion people globally and remains a leading cause of disability [67]. Although soft tissue mechanotherapies have been shown to provide pain relief [20], [21], [22], their mechanisms of action are still not fully understood, and no reliable quantitative method exists to classify tissue states in relation to pain. This study developed an optical approach to quantify soft tissue mobility using strain-based biomarkers, aiming to bridge this gap and offer a tool that complements clinicians’ existing workflows. Our primary hypothesis, that these biomarkers would be sensitive enough to detect variations in tissue mobility within individuals, was supported by our findings. While no overall trends were found at the population level based on body side, individual-level analysis identified significant bilateral differences in nine participants, with a high correlation to self-reported pain levels in the majority of these cases. Furthermore, systematic directional trends suggest that anatomical differences influence soft tissue mobility, with inferior pulls generally producing greater tissue glide and superior pulls eliciting greater tissue deformation.

The results establish a link between self-reported pain and quantitative biomarkers of soft tissue mobility, advancing the clinical potential of these biomarkers for more precise and consistent assessments of myofascial pain. The ability to objectively measure soft tissue mobility offers a pathway toward evidence-based, personalized therapeutic interventions, improving clinical decision-making. Moreover, by providing a robust tool for tissue assessment, this study opens new directions for future research into the relationship between tissue properties, pain perception, and clinical outcomes. This research not only helps to address the challenges associated with subjective pain measurement but also lays the foundation for more targeted treatments, marking an important step toward precision medicine in musculoskeletal care.

### 4.1. Bilateral mobility differences align with pain

Nine participants were identified as bilaterally different by DIC, eight of whom reported discrepancies in bilateral pain ratings (Table 2; Fig. 7). For instance, participant 13 showed significant bilateral differences in 14 out of 22 biomarkers (Fig. 6). Specifically, tissue in this participant’s left anatomy, when pulled superiorly, achieved lower values of maximum pull, maximum far region displacement, and pull propagation, compared to the right anatomy (Supp. Fig. 4(c), (g), and (i)). However, the left anatomy tissue simultaneously exhibited higher values of maximum tensile strain, strain at 5 mm, and strain-to-pull at maximum pull (Supp. Fig. 4(a), (q), and (s)). These results indicate that the left anatomy tissue glided less and deformed more than the right anatomy tissue, suggesting that participant 13 may have underlying myofascial impairment of the left anatomy. Healthy tissue typically allows the skin and subcutaneous layers to glide over deeper structures [68], resulting in higher values for maximum pull and pull propagation. In contrast, tissue impaired with myofascial trigger points restricts tissue glide [7], [8], necessitating greater deformation to accommodate the applied force. While this case highlights a potential link between tissue behavior and underlying myofascial impairment, broader physiological factors may also play a role. For example, although BMI collected during intake showed no significant correlation with biomarkers (*p* > 0.25), this does not preclude the influence of body composition on tissue properties, which warrants further investigation in larger, more diverse samples.

Interestingly, bilateral differences were at times confined to a single pull direction. For example, P8 had eight biomarkers exceeding the threshold in the superior direction but none in the inferior direction (Table 2). This finding could help localize MTrPs, as a trigger point higher in the cervicothoracic region might restrict mobility during superior pulls but have minimal impact during inferior pulls. Indeed, previous studies report that MTrP response and measured stiffness decreases with increasing longitudinal distance from applied force [69].

Overall, the DIC methods showed an 84.2% alignment with self-reported pain discrepancies (Fig. 7), highlighting their effectiveness compared to other quantitative methods where the relationship between pain intensity and muscle properties remains unclear [70], [71], [72], [73], [74]. While no systematic bilateral trends were observed (Fig. 4(i)-(l)), this absence may suggest a predisposition for spinal symmetry in the cervicothoracic region [75], [76]. However, lifestyle factors such as physical injury, muscle overuse, and poor ergonomics can disrupt this symmetry, likely contributing to the observed asymmetries and myofascial pain in some participants [69].

### 4.2. Directional differences highlight anatomical asymmetries

Systematic directional trends in tissue mobility were observed at the population level and prominently measured in sixteen out of nineteen participants when assessed individually. For example, biomarkers related to tissue glide, such as maximum pull (Fig. 4(e)), maximum far region displacement (Fig. 4(f)), and pull propagation (Supp. Fig. 2(h)), were higher in the inferior direction. In contrast, biomarkers measuring deformation under equivalent loading conditions, including ramp-on strain gradient, strain at 5 mm, and strain-to-pull at maximum pull (Supp. Fig. 2(j), (l), and (m)), were higher in the superior direction. These widespread directional distinctions likely reflect differences in anatomical structure between the pull locations. Indeed, the cervicothoracic region’s complex tissue architecture, influenced by overlapping muscles such as the trapezius, rhomboids, and erector spinae, may exhibit anisotropic behavior. Furthermore, Langer’s lines, which align with underlying muscle fibers and fascia [77], vary in orientation across different body sites and during muscle contractions [78], [79]. For instance, Langer’s lines are predominantly horizontal in the upper cervicothoracic region but become more oblique in the middle cervicothoracic region [77]. Testing perpendicular to Langer’s lines typically shows lower ultimate tensile strength, as collagen fibers are coiled and gradually extend before reaching maximum stiffness [75]. In contrast, testing parallel to these lines results in maximum stiffness at lower stretch ratios due to initially taut collagen fibers with less capacity for further expansion.

Although directional distinctions were observed in sixteen participants, some individuals deviated from the population-level trends. For instance, one participant (P17) exhibited substantially higher maximum pull in the superior direction on both the left (superior: 46.3 mm; inferior: 23.4 mm) and right sides (superior: 39.8 mm; inferior: 22.4 mm) (Supp. Fig. 3(c) and (d)). Furthermore, P17 also demonstrated lower tensile strain-to-pull ratios at maximum pull in the superior direction on the left (superior: 0.04 1/cm; inferior: 0.07 1/cm) and right sides (superior: 0.04 1/cm; inferior: 0.07 1/cm) (Supp. Fig. 3(s) and (t)). While no bilateral pain discrepancy was reported in the cervicothoracic region for this participant, these results may indicate restricted mobility in other body regions. The interconnected nature of the fascial system, specifically the superficial back line (SBL), means that tension in the lower back can be transmitted upwards through the thoracolumbar fascia and erector spinae muscles, affecting the entire back [80].

### 4.3. In vivo tissue analysis by 3D-DIC

In this study we developed a non-invasive 3D-DIC method to dynamically capture clinician-performed STM stretch in the cervicothoracic region. We aimed to objectively measure and quantitatively characterize clinically relevant aspects of myofascial mobility, such as the spatial extent and rate of tissue displacement during clinician-applied loading. We found that approximately 62.7% of the clinician-applied maximum pull (30.3 mm; Fig. 4(a)) propagated across the tracked region (19.0 mm; Fig. 4(b)), while the remaining 37.3% dissipated through the tissue as it deformed. Tissue deformation was measured as principal strains, the magnitudes of which align closely with prior studies using DIC during passive and active stretch. In particular, Maiti et al. captured mean principal strains of 26.1% in tension and −8.4% in compression in the volar forearm during active extension from 90° to 180° [29]. Similarly, the median strain measurements in this study, though passive rather than active, ranged from 0.22 (22%) in tension (Fig. 4(c)) to −0.14 (−14%) in compression (Fig. 4(d)). Furthermore, Solav et al. measured principal stretches of 80% - 120% in the calf region of the lower leg while a subject performed ankle plantarflexion [56]. To draw a comparison to our findings, we can convert our principal strains (ε_i_) to principal stretch (λ_i_) values using (4) [81].

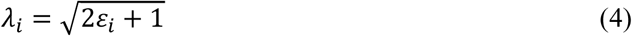

In doing so, we find comparable results with 120% tensile and 85% compressive principal stretch. That stated, skin strain measured in literature can vary widely based on experimental procedure, body locations, and participant characteristics.

While previous studies have used DIC methods to characterize the mechanical properties of skin [82], [83], [84], investigate tactile afferent responses [29], and assess internal tissue movements [85], [86], this is, to our knowledge, the first application of these methods for assessing passive skin displacement during soft tissue manipulation, which may be influenced by underlying myofascial mobility. A key advantage of this approach is that it allows for hands-on treatment, preserving the psychosocial benefits of human touch [53]. Furthermore, the use of open-source software and low-cost hardware, combined with the novel semi-permanent speckling method, enables patient-friendly, reproducible, and longitudinal assessments of treatment effects.

### 4.4. Challenges to objective myofascial measurements

Accurately characterizing myofascial tissue states remains a challenge. Specifically, the complex interplay between patient perceptions, clinician impressions, and quantitative metrics makes it difficult to universally identify an objective ground truth. For example, clinician assessments can be influenced by their experience, style, expectations, and participant feedback, which may introduce an ‘observer effect’ [87]. To minimize bias due to clinician experience, we opted to work with a single clinician. To further mitigate the ‘observer effect’, we prioritized participant pain ratings as a more direct and uninfluenced measure of tissue mobility. Another complicating factor are differences in pain phenotypes that may be associated with myofascial dysfunction. Although inherently subjective and shaped by both physiological and psychological factors, pain remains a central focus of many manual therapies and offers valuable insights [50], [51].

While methods like algometry, myotonometry, and shear wave elastography have been explored as alternatives to pain ratings, they often lack consistency and reliability. Algometry measures tissue pain pressure thresholds but varies across body sites and practitioners, impacting its reliability for identifying myofascial trigger points [88]. Myotonometers have captured correlation between muscle stiffness and pain [89], but are constrained by their small measurable area and sensitivity to muscle contraction, leading to inconsistent results [90]. Shear wave elastography provides detailed images of tissue stiffness but suffers from operator and device variability, lack of standardized protocols, and reduced accuracy in anisotropic tissues due to transducer angle effects [27]. For instance, one study found no significant differences between latent and active myofascial trigger points despite pain pressure threshold differences, indicating potential disconnects with clinical severity indicators like pain intensity [91]. These limitations underscore the need for alternative methods to provide a more comprehensive assessment of tissue mechanics.

### 4.5. Future directions and clinical translation

In this study, we adopted an observational framework that allowed an experienced clinician to perform soft tissue manipulation without imposed constraints, while maintaining consistency through a single practitioner and standardized data processing. Rather than estimating absolute force values or deriving ground-truth mechanical properties, our goal was to detect relative differences in tissue behavior across sides and directions and explore their relationship to participant pain. Although our cohort of participants was modest in size, we successfully validated our exploratory methodologies prior to conducting to a larger-scale clinical trial. The absence of an objective ground truth and the potential for multiple sources of bias necessitated a limited case study design until the efficacy of the methods was confirmed, consistent with prior exploratory studies evaluating manual therapies [27], [52], [92]. Importantly, our initial objective was to observe and quantify the STM maneuver as it is performed in practice—unblinded, tactile, and clinician-driven—because this approach is already used effectively in clinical assessment of myofascial pain. Now that the process has been validated using quantitative surface tracking, future studies will refine the protocol to enhance measurement accuracy and rigorously assess validity and reliability.

In the next phase of research, we plan to introduce a second clinician to assess inter-clinician variability and to incorporate independent clinical evaluations of tissue state as an unbiased comparator to pain ratings. We are also interested in exploring inter-clinician variability in greater depth, examining how the maneuver may vary across practitioners. Furthermore, we are now well-positioned to evaluate therapeutic effects by applying the same methodology before and after soft tissue intervention, while preserving the essential features of the maneuver that make it effective in real-world practice.

## 5. Conclusions

This study developed an optical method to derive skin surface biomarkers for characterizing soft tissue mobility, wherein a DIC based approach was customized for use in the skin, with particular focus on measuring tissue glide and deformation. In a study of nineteen participants, DIC-identified bilateral differences aligned with self-reported pain in 84.2% of cases, demonstrating the robust potential of these methods to objectively and reliably quantify soft tissue mobility. The development of sensitive biomarkers that maintain direct skin contact between clinician and patient, without altering proven clinical techniques, may enhance the precision of affective touch therapies for myofascial pain, providing an objective, evidence-based approach to assessing and treating this condition, which affects over 250 million Americans.

## Data Availability

All data produced in the present study are available upon reasonable request to the authors.

## CRediT authorship contribution statement

**Anika Kao:** Conceptualization, data curation, formal analysis, investigation, methodology, software, validation, visualization, writing – original draft, writing – review and editing. **Terry Loghmani:** Conceptualization, funding acquisition, investigation, methodology, writing – review and editing. **Gregory Gerling:** Conceptualization, funding acquisition, methodology, supervision, writing – original draft, writing – review and editing.

## References

[1] T. Sundberg, H. Cramer, D. Sibbritt, J. Adams, and R. Lauche, “Prevalence, patterns, and predictors of massage practitioner utilization: Results of a US nationally representative survey,” Musculoskeletal Science and Practice, vol. 32, pp. 31–37, Dec. 2017.

[2] R. Adams, B. White, and C. Beckett, “The Effects of Massage Therapy on Pain Management in the Acute Care Setting,” Int J Ther Massage Bodywork, vol. 3, no. 1, pp. 4–11, Mar. 2010.

[3] J. W. Lucas and I. Sohi, “Chronic Pain and High-impact Chronic Pain Among U.S. Adults, 2023,” National Center for Health Statistics (U.S.), Hyattsville, MD, Nov. 2024.

[4] R. L. Nahin, A. Rhee, and B. Stussman, “Use of Complementary Health Approaches Overall and for Pain Management by US Adults,” JAMA, vol. 331, no. 7, pp. 613–615, Feb. 2024.

[5] Headache Classification Committee of the International Headache Society (IHS), “The International Classification of Headache Disorders, 3rd edition (beta version),” Cephalalgia, vol. 33, no. 9, pp. 629–808, Jul. 2013.

[6] GBD 2015 Neurological Disorders Collaborator Group, “Global, regional, and national burden of neurological disorders during 1990-2015: a systematic analysis for the Global Burden of Disease Study 2015,” Lancet Neurol, vol. 16, no. 11, pp. 877–897, Nov. 2017.

[7] D. G. Simons, “Clinical and Etiological Update of Myofascial Pain from Trigger Points,” Journal of Musculoskeletal Pain, vol. 4, no. 1–2, pp. 93–122, Jan. 1996.

[8] T. P. Do, G. F. Heldarskard, L. T. Kolding, J. Hvedstrup, and H. W. Schytz, “Myofascial trigger points in migraine and tension-type headache,” J Headache Pain, vol. 19, no. 1, p. 84, Sep. 2018.

[9] D. G. Simons, J. G. Travell, and L. S. Simons, Travell & Simons’ myofascial pain and dysfunction: the trigger point manual, 2nd ed. Baltimore: Williams & Wilkins, 1999.

[10] A. Streďanská, D. Nečas, M. Vrbka, J. Suchánek, J. Matonohová, E. Toropitsyn, M. Hartl, I. Křupka, and K. Nešporová, “Understanding frictional behavior in fascia tissues through tribological modeling and material substitution,” Journal of the Mechanical Behavior of Biomedical Materials, vol. 155, p. 106566, Jul. 2024.

[11] C. H. Karels, W. Polling, S. M. A. Bierma-Zeinstra, A. Burdorf, A. P. Verhagen, and B. W. Koes, “Treatment of arm, neck, and/or shoulder complaints in physical therapy practice,” Spine (Phila Pa 1976), vol. 31, no. 17, pp. E584–589, Aug. 2006.

[12] S. W. Cheatham, R. Baker, and E. Kreiswirth, “Instrument Assisted Soft Tissue Mobilization: A commentary on clinical practice guidelines for rehabilitation professionals,” Int J Sports Phys Ther, vol. 14, no. 4, pp. 670–682, Jul. 2019.

[13] “AMTA,” American Massage Therapy Association. Accessed: Feb. 08, 2022. [Online]. Available: https://www.amtamassage.org/

[14] T. Loghmani M and S. Bane, “Instrument-assisted Soft Tissue Manipulation: Evidence for its Emerging Efficacy,” J Nov Physiother, vol. s3, 2016.

[15] A. Bhattacharjee, S. Anwar, S. Chien, and M. T. Loghmani, “A Handheld Quantifiable Soft Tissue Manipulation Device for Tracking Real-Time Dispersive Force-Motion Patterns to Characterize Manual Therapy Treatment,” IEEE Trans Biomed Eng, vol. PP, Nov. 2022.

[16] R. I. Cantu, A. J. Grodin, and R. W. Stanborough, Myofascial manipulation: Theory and clinical application, 3rd ed. in Aspen series in physical therapy. Austin, Texas: PRO-ED, Incorporated, 2012.

[17] D. Morgan, “Principles of Soft Tissue Treatment,” Journal of Manual & Manipulative Therapy, vol. 2, no. 2, pp. 63–65, Jan. 1994.

[18] “American Physical Therapy Association,” APTA. Accessed: Feb. 08, 2022. [Online]. Available: https://www.apta.org/

[19] D. L. Keter, J. A. Bent, J. E. Bialosky, C. A. Courtney, J. E. Esteves, M. Funabashi, S. J. Howarth, H. S. Injeyan, A. M. Mazzieri, C. Glissmann Nim, and C. E. Cook, “An international consensus on gaps in mechanisms of forced-based manipulation research: findings from a nominal group technique,” J Man Manip Ther, vol. 32, no. 1, pp. 111–117, Feb. 2024.

[20] D. T. Gulick, “Instrument-assisted soft tissue mobilization increases myofascial trigger point pain threshold,” J Bodyw Mov Ther, vol. 22, no. 2, pp. 341–345, Apr. 2018.

[21] C. Cumplido-Trasmonte, P. Fernández-González, I. M. Alguacil-Diego, and F. Molina-Rueda, “Manual therapy in adults with tension-type headache: A systematic review,” Neurologia (Engl Ed), vol. 36, no. 7, pp. 537–547, Sep. 2021.

[22] G. Jull, P. Trott, H. Potter, G. Zito, K. Niere, D. Shirley, J. Emberson, I. Marschner, and C. Richardson, “A Randomized Controlled Trial of Exercise and Manipulative Therapy for Cervicogenic Headache,” Spine, vol. 27, no. 17, p. 1835, Sep. 2002.

[23] S. Engell, J. J. Triano, J. R. Fox, H. M. Langevin, and E. E. Konofagou, “Differential displacement of soft tissue layers from manual therapy loading,” Clinical Biomechanics, vol. 33, pp. 66–72, Mar. 2016.

[24] J. Vappou, C. Maleke, and E. E. Konofagou, “Quantitative viscoelastic parameters measured by harmonic motion imaging,” Phys Med Biol, vol. 54, no. 11, pp. 3579–3594, Jun. 2009.

[25] D. A. Kumbhare, A. H. Elzibak, and M. D. Noseworthy, “Assessment of Myofascial Trigger Points Using Ultrasound,” Am J Phys Med Rehabil, vol. 95, no. 1, pp. 72–80, Jan. 2016.

[26] A. Romano, D. Staber, A. Grimm, C. Kronlage, and J. Marquetand, “Limitations of Muscle Ultrasound Shear Wave Elastography for Clinical Routine-Positioning and Muscle Selection,” Sensors (Basel), vol. 21, no. 24, p. 8490, Dec. 2021.

[27] A. S. Vuorenmaa, E. M. K. Siitama, and K. S. Mäkelä, “Inter-operator and inter-device reproducibility of shear wave elastography in healthy muscle tissues,” J Appl Clin Med Phys, vol. 23, no. 9, p. e13717, Sep. 2022.

[28] X. Hu, R. Maiti, X. Liu, L. C. Gerhardt, Z. S. Lee, R. Byers, S. E. Franklin, R. Lewis, S. J. Matcher, and M. J. Carré, “Skin surface and sub-surface strain and deformation imaging using optical coherence tomography and digital image correlation,” in Optical Elastography and Tissue Biomechanics III, International Society for Optics and Photonics, Mar. 2016, p. 971016.

[29] R. Maiti, L.-C. Gerhardt, Z. S. Lee, R. A. Byers, D. Woods, J. A. Sanz-Herrera, S. E. Franklin, R. Lewis, S. J. Matcher, and M. J. Carré, “In vivo measurement of skin surface strain and sub-surface layer deformation induced by natural tissue stretching,” Journal of the Mechanical Behavior of Biomedical Materials, vol. 62, pp. 556–569, Sep. 2016.

[30] Y. Yu, H. Wang, P. Chen, Y. Zhang, Z. Guo, and R. Liang, “A New Approach to External and Internal Fingerprint Registration With Multisensor Difference Minimization,” IEEE Transactions on Biometrics, Behavior, and Identity Science, vol. 2, no. 4, pp. 363–376, Oct. 2020.

[31] L. Chuang, C. Wu, and K. Lin, “Reliability, Validity, and Responsiveness of Myotonometric Measurement of Muscle Tone, Elasticity, and Stiffness in Patients With Stroke,” Archives of Physical Medicine and Rehabilitation, vol. 93, no. 3, pp. 532–540, Mar. 2012.

[32] C. M. Kerins, S. D. Moore, T. A. Butterfield, P. O. McKeon, and T. L. Uhl, “RELIABILITY OF THE MYOTONOMETER FOR ASSESSMENT OF POSTERIOR SHOULDER TIGHTNESS,” International Journal of Sports Physical Therapy, vol. 8, no. 3, p. 248, Jun. 2013.

[33] C. T. Leonard, W. P. Deshner, J. W. Romo, E. S. Suoja, S. C. Fehrer, and E. L. Mikhailenok, “Myotonometer Intra- and Interrater Reliabilities1,” Archives of Physical Medicine and Rehabilitation, vol. 84, no. 6, pp. 928–932, Jun. 2003.

[34] R. Gulati and M. Srinivasan, “Human fingerpad under indentation I: static and dynamic force response,” undefined, 1995. Accessed: Mar. 31, 2022. [Online]. Available: https://www.semanticscholar.org/paper/Human-fingerpad-under-indentation-I%3A-static-and-Gulati-Srinivasan/7d258c6ac431b582a2c13c1e5247505c6369807e

[35] Y. Wang and G. J. Gerling, “Computational Modeling Reinforces that Proprioceptive Cues May Augment Compliance Discrimination When Elasticity Is Decoupled from Radius of Curvature,” in Haptics: Neuroscience, Devices, Modeling, and Applications: 9th International Conference, EuroHaptics 2014, Versailles, France, June 24-26, 2014, Proceedings, Part II, M. Auvray and C. Duriez, Eds., Berlin, Heidelberg: Springer Berlin Heidelberg, 2014, pp. 360–368. [Online]. Available: 10.1007/978-3-662-44196-1_44

[36] C. Xu, H. He, S. C. Hauser, and G. J. Gerling, “Tactile Exploration Strategies With Natural Compliant Objects Elicit Virtual Stiffness Cues,” IEEE Transactions on Haptics, vol. 13, no. 1, pp. 4–10, Jan. 2020.

[37] C. Shanley, Q. J. Wang, and B. Livingston, “Approach for contact medical device development via integrated testing, skeletal muscle modeling, and finite element analysis,” Journal of the Mechanical Behavior of Biomedical Materials, vol. 155, p. 106541, Jul. 2024.

[38] S. C. Hauser, S. S. Nagi, S. McIntyre, A. Israr, H. Olausson, and G. J. Gerling, “From Human-to-Human Touch to Peripheral Nerve Responses,” in 2019 IEEE World Haptics Conference (WHC), Jul. 2019, pp. 592–597.

[39] S. Xu, C. Xu, S. McIntyre, H. Olausson, and G. J. Gerling, “3D Visual Tracking to Quantify Physical Contact Interactions in Human-to-Human Touch,” Frontiers in Physiology, vol. 13, 2022. Accessed: Feb. 28, 2023. [Online]. Available: https://www.frontiersin.org/articles/10.3389/fphys.2022.841938

[40] C. Lo, S. T. Chu, T. B. Penney, and A. Schirmer, “3D Hand-Motion Tracking and Bottom-Up Classification Sheds Light on the Physical Properties of Gentle Stroking,” Neuroscience, Sep. 2020.

[41] S. Sridhar, F. Mueller, A. Oulasvirta, and C. Theobalt, “Fast and robust hand tracking using detection-guided optimization,” in 2015 IEEE Conference on Computer Vision and Pattern Recognition (CVPR), Jun. 2015, pp. 3213–3221.

[42] E. K. Vonstad, X. Su, B. Vereijken, K. Bach, and J. H. Nilsen, “Comparison of a Deep Learning-Based Pose Estimation System to Marker-Based and Kinect Systems in Exergaming for Balance Training,” Sensors, vol. 20, no. 23, Art. no. 23, Jan. 2020.

[43] B. Li, S. Hauser, and G. J. Gerling, “Identifying 3-D spatiotemporal skin deformation cues evoked in interacting with compliant elastic surfaces,” in 2020 IEEE Haptics Symposium (HAPTICS), Mar. 2020, pp. 35–40.

[44] A. R. Kao, C. Xu, and G. J. Gerling, “Using Digital Image Correlation to Quantify Skin Deformation With Von Frey Monofilaments,” IEEE Transactions on Haptics, vol. 15, no. 1, pp. 26–31, Jan. 2022.

[45] A. R. Kao, Z. T. Landsman, G. J. Gerling, and M. T. Loghmani, “Optical Measurements of the Skin Surface to Infer Bilateral Distinctions in Myofascial Tissue Stiffness,” in 2023 IEEE World Haptics Conference (WHC), Jul. 2023, pp. 244–251.

[46] D. Solav, K. M. Moerman, A. M. Jaeger, and H. M. Herr, “A Framework for Measuring the Time-Varying Shape and Full-Field Deformation of Residual Limbs Using 3-D Digital Image Correlation,” IEEE Transactions on Biomedical Engineering, vol. 66, no. 10, pp. 2740–2752, Oct. 2019.

[47] Z. S. Lee, R. Maiti, M. J. Carré, and R. Lewis, “Morphology of a human finger pad during sliding against a grooved plate: A pilot study,” Biotribology, vol. 21, p. 100114, Mar. 2020.

[48] Z. Xu, J. D. Cruz, C. Fthenakis, and C. Saliou, “A novel method to measure skin mechanical properties with three-dimensional digital image correlation,” Skin Research and Technology, vol. 25, no. 1, pp. 60–67, 2019.

[49] S. R. Crossland, H. J. Siddle, P. Culmer, and C. L. Brockett, “A plantar surface shear strain methodology utilising Digital Image Correlation,” Journal of the Mechanical Behavior of Biomedical Materials, vol. 136, p. 105482, Dec. 2022.

[50] C. Bernal-Utrera, J. J. Gonzalez-Gerez, E. Anarte-Lazo, and C. Rodriguez-Blanco, “Manual therapy versus therapeutic exercise in non-specific chronic neck pain: a randomized controlled trial,” Trials, vol. 21, no. 1, p. 682, Jul. 2020.

[51] J. Castro, L. Correia, B. de S. Donato, B. Arruda, F. Agulhari, M. J. Pellegrini, F. T. C. Belache, C. P. de Souza, J. Fernandez, L. A. C. Nogueira, F. J. J. Reis, A. de S. Ferreira, and N. Meziat-Filho, “Cognitive functional therapy compared with core exercise and manual therapy in patients with chronic low back pain: randomised controlled trial,” Pain, vol. 163, no. 12, pp. 2430–2437, Dec. 2022.

[52] Y. Nadal-Nicolás, J. Á. Rubio-Arias, M. Martínez-Olcina, C. Reche-García, M. Hernández-García, and A. Martínez-Rodríguez, “Effects of Manual Therapy on Fatigue, Pain, and Psychological Aspects in Women with Fibromyalgia,” Int J Environ Res Public Health, vol. 17, no. 12, p. 4611, Jun. 2020.

[53] M. Rezaei, S. S. Nagi, C. Xu, S. McIntyre, H. Olausson, and G. J. Gerling, “Thin Films on the Skin, but not Frictional Agents, Attenuate the Percept of Pleasantness to Brushed Stimuli,” in 2021 IEEE World Haptics Conference (WHC), Jul. 2021, pp. 49–54.

[54] C. Xu, Y. Wang, and G. J. Gerling, “An elasticity-curvature illusion decouples cutaneous and proprioceptive cues in active exploration of soft objects,” PLOS Computational Biology, vol. 17, no. 3, p. e1008848, Mar. 2021.

[55] S. C. Hauser and G. J. Gerling, “Force-rate Cues Reduce Object Deformation Necessary to Discriminate Compliances Harder than the Skin,” IEEE Trans Haptics, vol. 11, no. 2, pp. 232–240, 2018.

[56] D. Solav, K. M. Moerman, A. M. Jaeger, K. Genovese, and H. M. Herr, “MultiDIC: An Open-Source Toolbox for Multi-View 3D Digital Image Correlation,” IEEE Access, vol. 6, pp. 30520–30535, 2018.

[57] J. Blaber, B. Adair, and A. Antoniou, “Ncorr: Open-Source 2D Digital Image Correlation Matlab Software,” Exp Mech, vol. 55, no. 6, pp. 1105–1122, Jul. 2015.

[58] O. Levano Blanch, D. Lunt, G. J. Baxter, and M. Jackson, “Deformation Behaviour of a FAST Diffusion Bond Processed from Dissimilar Titanium Alloy Powders,” Metall Mater Trans A, vol. 52, no. 7, pp. 3064–3082, Jul. 2021.

[59] A. Yoshiko, R. Ando, and H. Akima, “Passive muscle stiffness is correlated with the intramuscular adipose tissue in young individuals,” Eur J Appl Physiol, vol. 123, no. 5, pp. 1081–1090, May 2023.

[60] S. F. Eby, B. A. Cloud, J. E. Brandenburg, H. Giambini, P. Song, S. Chen, N. K. LeBrasseur, and K.-N. An, “Shear wave elastography of passive skeletal muscle stiffness: influences of sex and age throughout adulthood,” Clin Biomech (Bristol), vol. 30, no. 1, pp. 22–27, Jan. 2015.

[61] Y. Wang, K. L. Marshall, Y. Baba, E. A. Lumpkin, and G. J. Gerling, “Compressive Viscoelasticity of Freshly Excised Mouse Skin Is Dependent on Specimen Thickness, Strain Level and Rate,” PLOS ONE, vol. 10, no. 3, p. e0120897, Mar. 2015.

[62] M. Pourahmadi, J. Dommerholt, C. Fernández-de-Las-Peñas, B. W. Koes, M. A. Mohseni-Bandpei, M. A. Mansournia, S. Delavari, A. Keshtkar, and M. Bahramian, “Dry Needling for the Treatment of Tension-Type, Cervicogenic, or Migraine Headaches: A Systematic Review and Meta-Analysis,” Physical Therapy, vol. 101, no. 5, p. pzab068, May 2021.

[63] M. J. Kelley, M. A. Shaffer, J. E. Kuhn, L. A. Michener, A. L. Seitz, T. L. Uhl, J. J. Godges, and P. McClure, “Shoulder Pain and Mobility Deficits: Adhesive Capsulitis,” Journal of Orthopaedic & Sports Physical Therapy, vol. 43, no. 5, pp. A1–A31, May 2013.

[64] J. W. DeVocht, J. G. Pickar, and D. G. Wilder, “Spinal Manipulation Alters Electromyographic Activity of Paraspinal Muscles: A Descriptive Study,” Journal of Manipulative and Physiological Therapeutics, vol. 28, no. 7, pp. 465–471, Sep. 2005.

[65] J. Hvedstrup, L. T. Kolding, M. Ashina, and H. W. Schytz, “Increased neck muscle stiffness in migraine patients with ictal neck pain: A shear wave elastography study,” Cephalalgia, vol. 40, no. 6, pp. 565–574, May 2020.

[66] L. Proulx, K. Brizzolara, M. Thompson, S. Wang-Price, P. Rodriguez, and S. Koppenhaver, “Women with Chronic Pelvic Pain Demonstrate Increased Lumbopelvic Muscle Stiffness Compared to Asymptomatic Controls,” J Womens Health (Larchmt), vol. 32, no. 2, pp. 239–247, Feb. 2023.

[67] A. Cieza, K. Causey, K. Kamenov, S. W. Hanson, S. Chatterji, and T. Vos, “Global estimates of the need for rehabilitation based on the Global Burden of Disease study 2019: a systematic analysis for the Global Burden of Disease Study 2019,” Lancet, vol. 396, no. 10267, pp. 2006–2017, Dec. 2021.

[68] H. M. Langevin, “Fascia Mobility, Proprioception, and Myofascial Pain,” Life (Basel), vol. 11, no. 7, p. 668, Jul. 2021.

[69] P. Tsai, J. Edison, C. Wang, J. Sefton, K. Q. Manning, and M. W. Gramlich, “Myofascial trigger point (MTrP) size and elasticity properties can be used to differentiate characteristics of MTrPs in lower back skeletal muscle,” Sci Rep, vol. 14, no. 1, p. 7562, Mar. 2024.

[70] K. Amris, A. Jespersen, and H. Bliddal, “Self-reported somatosensory symptoms of neuropathic pain in fibromyalgia and chronic widespread pain correlate with tender point count and pressure-pain thresholds,” PAIN, vol. 151, no. 3, p. 664, Dec. 2010.

[71] S. Mense and A. T. Masi, “Increased Muscle Tone as a Cause of Muscle Pain,” in Muscle Pain: Understanding the Mechanisms, S. Mense and R. D. Gerwin, Eds., Berlin, Heidelberg: Springer, 2010, pp. 207–249.

[72] C. Murillo, D. Falla, A. Rushton, A. Sanderson, and N. R. Heneghan, “Shear wave elastography investigation of multifidus stiffness in individuals with low back pain,” Journal of Electromyography and Kinesiology, vol. 47, pp. 19–24, Aug. 2019.

[73] W. L. A. Lo, Q. Yu, Y. Mao, W. Li, C. Hu, and L. Li, “Lumbar muscles biomechanical characteristics in young people with chronic spinal pain,” BMC Musculoskelet Disord, vol. 20, no. 1, p. 559, Nov. 2019.

[74] S. Taş, F. Korkusuz, and Z. Erden, “Neck Muscle Stiffness in Participants With and Without Chronic Neck Pain: A Shear-Wave Elastography Study,” Journal of Manipulative and Physiological Therapeutics, vol. 41, no. 7, pp. 580–588, Sep. 2018.

[75] M. Ottenio, D. Tran, A. Ní Annaidh, M. D. Gilchrist, and K. Bruyère, “Strain rate and anisotropy effects on the tensile failure characteristics of human skin,” J Mech Behav Biomed Mater, vol. 41, pp. 241–250, Jan. 2015.

[76] M. C. Corballis, “Bilaterally Symmetrical: To Be or Not to Be?,” Symmetry, vol. 12, no. 3, Art. no. 3, Mar. 2020.

[77] P. Agache, “Langer’s Lines,” in Agache’s Measuring the Skin, P. Humbert, H. Maibach, F. Fanian, and P. Agache, Eds., Cham: Springer International Publishing, 2016, pp. 1–6.

[78] C. J. Kraissl, “The selection of appropriate lines for elective surgical incisions,” Plast Reconstr Surg (1946), vol. 8, no. 1, pp. 1–28, Jul. 1951.

[79] D. Laiacona, J. M. Cohen, K. Coulon, Z. W. Lipsky, C. Maiorana, R. Boltyanskiy, E. R. Dufresne, and G. K. German, “Non-invasive in vivo quantification of human skin tension lines,” Acta Biomater, vol. 88, pp. 141–148, Apr. 2019.

[80] T. W. Myers, Anatomy Trains: Myofascial Meridians for Manual Therapists and Movement Professionals, 4th ed. Elsevier, 2021.

[81] J. W. Rudnicki, Fundamentals of Continuum Mechanics, 1st ed. Wiley, 2014. [Online]. Available: https://www.wiley.com/en-us/Fundamentals+of+Continuum+Mechanics-p-9781118479919

[82] S. Evans and C. Holt, “Measuring the Mechanical Properties of Human Skin in Vivo using Digital Image Correlation and Finite Element Modelling,” Journal of Strain Analysis for Engineering Design, vol. 44, Jul. 2009.

[83] P. Lakhani, K. K. Dwivedi, A. Parashar, and N. Kumar, “Non-Invasive in Vivo Quantification of Directional Dependent Variation in Mechanical Properties for Human Skin,” Front. Bioeng. Biotechnol., vol. 9, Oct. 2021.

[84] I. A. Staloff and M. Rafailovitch, “Measurement of skin stretch using digital image speckle correlation,” Skin Res Technol, vol. 14, no. 3, pp. 298–303, Aug. 2008.

[85] M. Hokka, N. Mirow, H. Nagel, M. Irqsusi, S. Vogt, and V.-T. Kuokkala, “In-vivo deformation measurements of the human heart by 3D Digital Image Correlation,” J Biomech, vol. 48, no. 10, pp. 2217–2220, Jul. 2015.

[86] Z. Chen, X. Shao, X. He, J. Wu, X. Xu, and J. Zhang, “Noninvasive, three-dimensional full-field body sensor for surface deformation monitoring of human body in vivo,” JBO, vol. 22, no. 9, p. 095001, Sep. 2017.

[87] S. E. Berger and A. T. Baria, “Assessing Pain Research: A Narrative Review of Emerging Pain Methods, Their Technosocial Implications, and Opportunities for Multidisciplinary Approaches,” Front Pain Res (Lausanne), vol. 3, p. 896276, 2022.

[88] W. Abu Taleb, A. Rehan Youssef, and A. Saleh, “The effectiveness of manual versus algometer pressure release techniques for treating active myofascial trigger points of the upper trapezius,” J Bodyw Mov Ther, vol. 20, no. 4, pp. 863–869, Oct. 2016.

[89] Z. Wu, Y. Zhu, W. Xu, J. Liang, Y. Guan, and X. Xu, “Analysis of Biomechanical Properties of the Lumbar Extensor Myofascia in Elderly Patients with Chronic Low Back Pain and That in Healthy People,” BioMed Research International, vol. 2020, no. 1, p. 7649157, 2020.

[90] Y. Lee, M. Kim, and H. Lee, “The Measurement of Stiffness for Major Muscles with Shear Wave Elastography and Myoton: A Quantitative Analysis Study,” Diagnostics (Basel), vol. 11, no. 3, p. 524, Mar. 2021.

[91] J. A. Valera-Calero, S. Sánchez-Jorge, J. Buffet-García, U. Varol, G. M. Gallego-Sendarrubias, and J. Álvarez-González, “Is Shear-Wave Elastography a Clinical Severity Indicator of Myofascial Pain Syndrome? An Observational Study,” Journal of Clinical Medicine, vol. 10, no. 13, Art. no. 13, Jan. 2021.

[92] G. E. Glassman, L. Dellalana, E. R. Tkaczyk, I. M. Esteve, J. Y. Huang, A. Cronin, J. R. Patrinely, S. Etemad, P. E. Assi, S. Ridner, A. J. Forte, S. A. Kassis, and G. Perdikis, “Measuring Biomechanical Properties Using a Noninvasive Myoton Device for Lymphedema Detection and Tracking: A Pilot Study,” Eplasty, vol. 22, p. e54, 2022.

