## Supplemental Figures for "Strain-based biomarkers at the skin surface differentiate asymmetries in soft tissue mobility associated with myofascial pain"

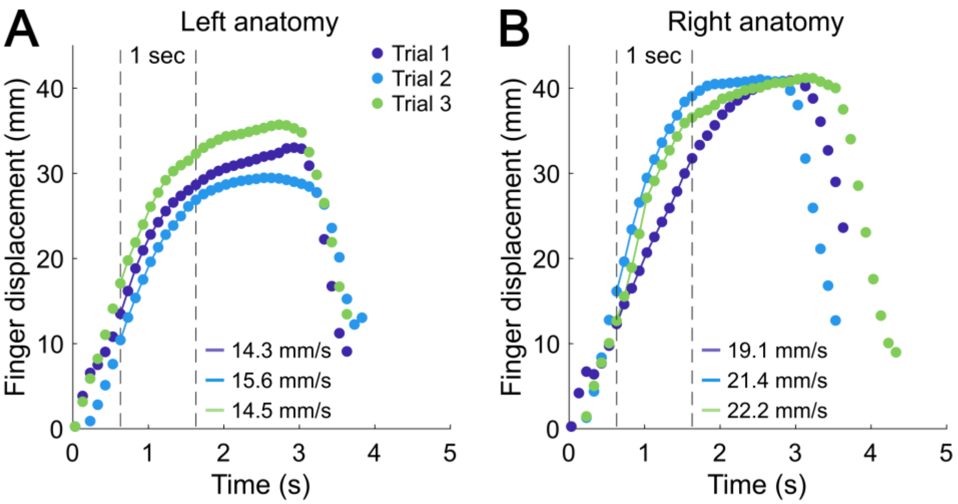


Supplemental Fig. 1. Clinician finger displacement over time during three repeated trials of STM stretch in the superior pull direction on the (a) left and (b) right anatomy for a representative participant. Each color denotes raw data from a separate trial, and the vertical dashed lines indicate the 1-second window used to compute average pull velocity, calculated as the mean derivative of displacement over this interval. Within each body side, pull velocity was highly consistent (left: 14.8 ± 0.7 mm/s; right: 20.9 ± 1.6 mm/s). Although velocities differed slightly between sides (6.1 mm/s), the influence of viscoelastic effects on strain output is expected to be negligible at such modest variations and timescales.

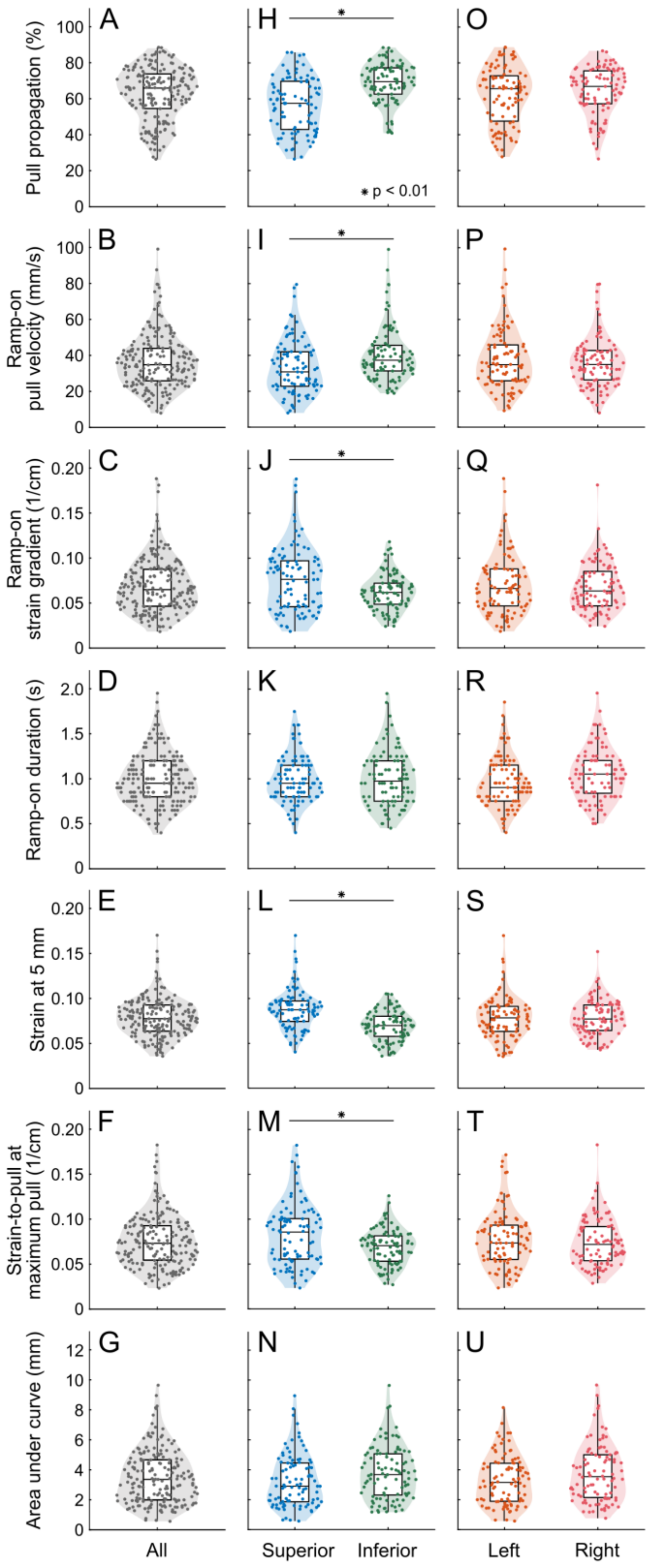


Supplemental Fig. 2. Aggregate population trends. (a)-(g) Biomarker distribution across all participants with pull directions and body sides aggregated. In (a) the pull propagation measured 65.9 ± 14.6% (median ± SD) while in (f) the strain-to-pull at maximum pull measured 0.07 ± 0.03 1/cm. (h)-(n) Aggregate data separated by pull direction and analyzed using linear mixed-effects models ($\alpha = 0.01$). Significant directional differences were observed in eight biomarkers (five shown: (h)-(j), (l)-(m)). Specifically, (h) pull propagation and (i) ramp-on pull velocity were greater in the inferior direction, while (j) ramp-on strain gradient, (l) strain at 5 mm, and (m) strain-to-pull at maximum pull were greater in the superior direction. (o)-(u) Aggregate data separated by body side and analyzed using linear mixed-effects models ($\alpha = 0.01$). No significant bilateral differences were found in any of the eleven biomarkers.

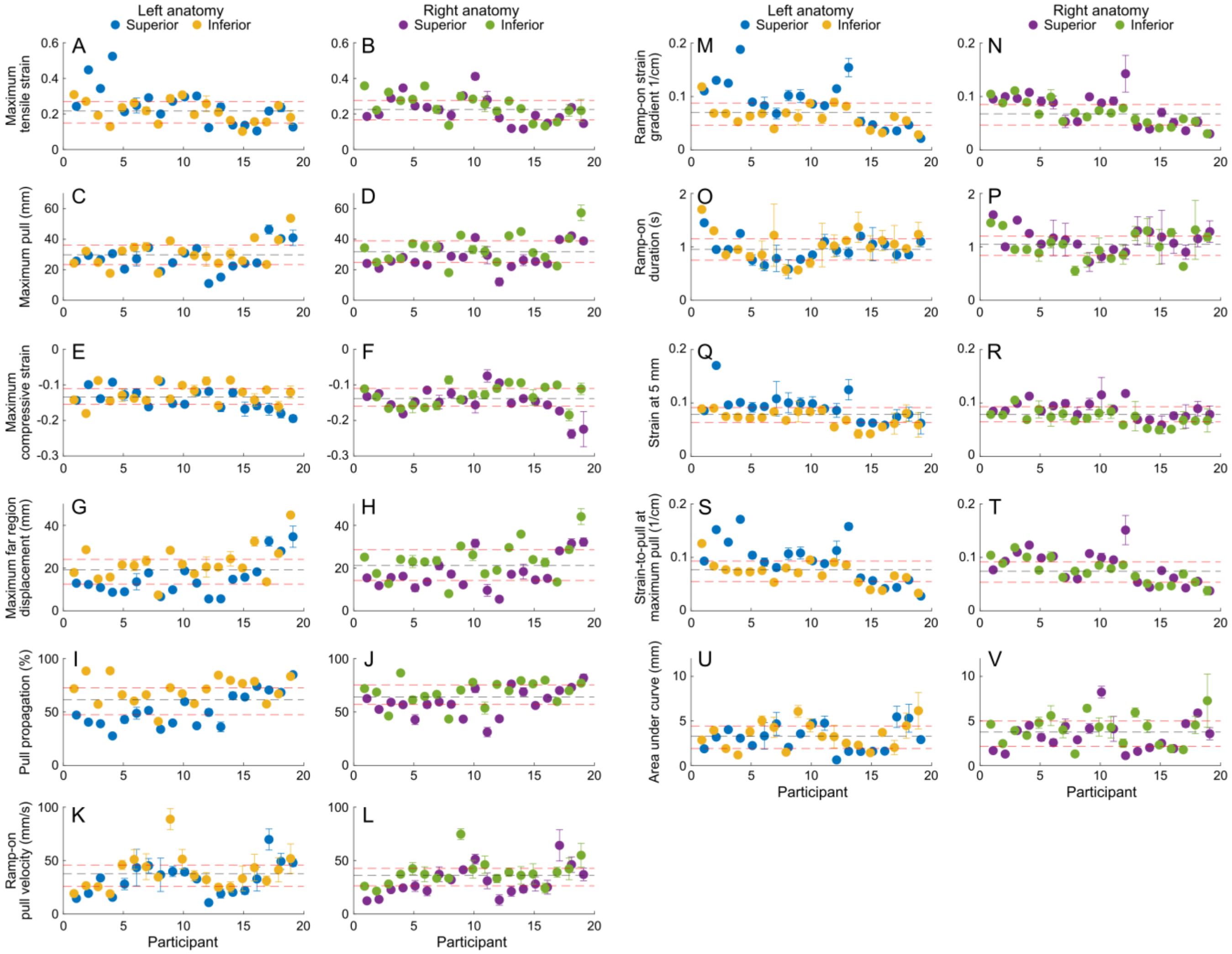


Supplemental Fig. 3. Directional comparisons at the individual level. Mean and standard deviation values for three trials per side and direction are marked for the left superior (blue), left inferior (yellow), right superior (purple), and right inferior (green) pulls. The population mean (black dashed line) and interquartile range (red dashed lines) are shown.

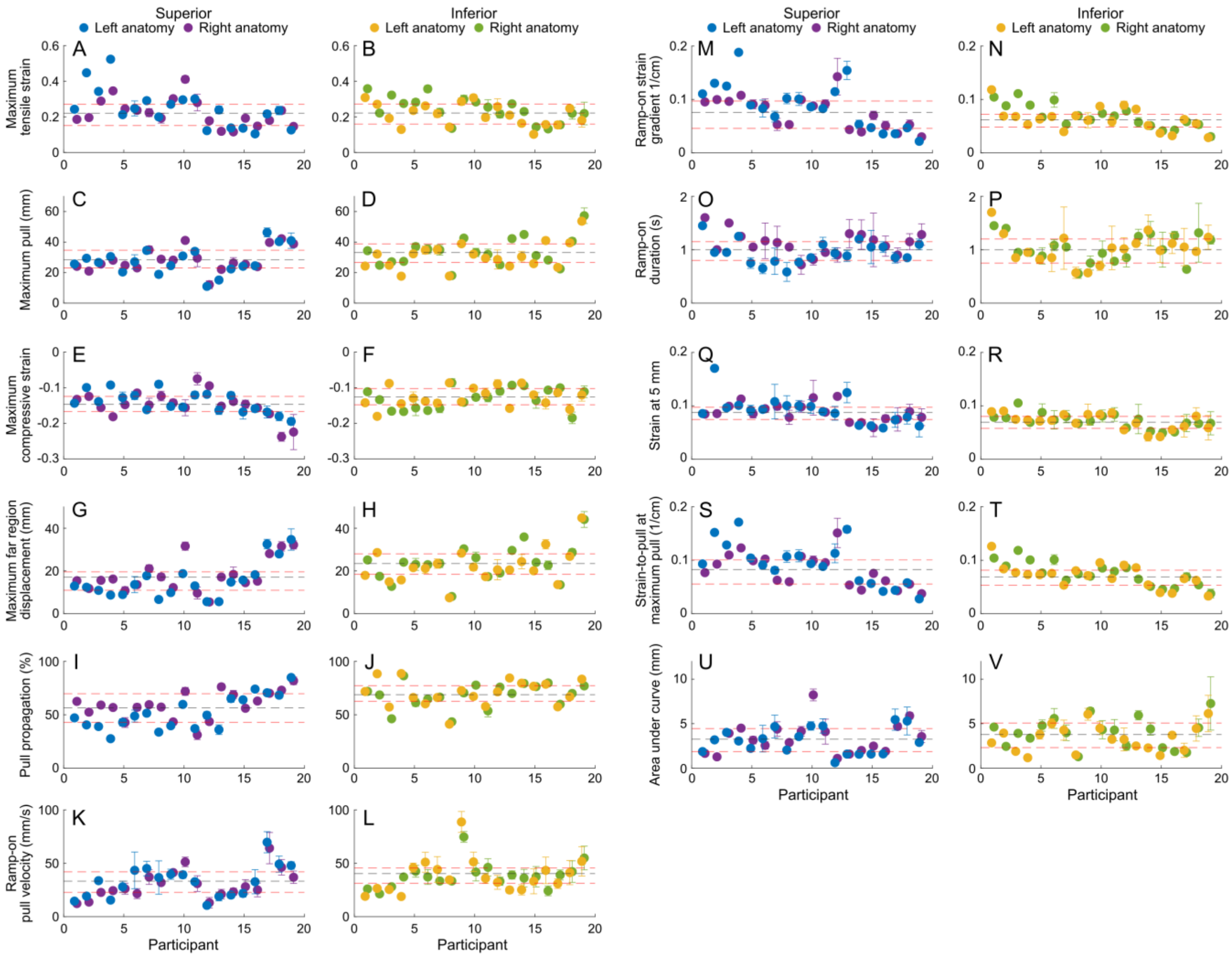


Supplemental Fig. 4. Bilateral comparisons at the individual level. Mean and standard deviation values for three trials per side and direction are marked for the superior left (blue), superior right (purple), inferior left (yellow), and inferior right (green) pulls. The population mean (black dashed line) and interquartile range (red dashed lines) are shown.
